# Deficiency of the minor spliceosome component U4atac snRNA secondarily results in ciliary defects

**DOI:** 10.1101/2021.12.12.21266616

**Authors:** Deepak Khatri, Audrey Putoux, Audric Cologne, Sophie Kaltenbach, Alicia Besson, Eloïse Bertiaux, Justine Guguin, Adèle Fendler, Marie A. Dupont, Clara Benoit-Pilven, Sarah Grotto, Lyse Ruaud, Caroline Michot, Martin Castelle, Agnès Guët, Laurent Guibaud, Virginie Hamel, Rémy Bordonné, Anne-Louise Leutenegger, Tania Attié-Bitach, Patrick Edery, Sylvie Mazoyer, Marion Delous

## Abstract

In the human genome, about 750 genes contain one intron excised by the minor spliceosome. This spliceosome comprises its own set of snRNAs, among which U4atac. Its non-coding gene, *RNU4ATAC*, has been found mutated in Taybi-Linder (MOPD1/TALS), Roifman (RFMN) and Lowry-Wood syndromes (LWS). These rare developmental disorders, whose physiopathological mechanisms remain unsolved, associate ante- and post-natal growth retardation, microcephaly, skeletal dysplasia, intellectual disability, retinal dystrophy and immunodeficiency. Here, we report a homozygous *RNU4ATAC* mutation in the Stem II domain, n.16G>A, in two unrelated patients presenting with both typical traits of the Joubert syndrome (JBTS), a well-characterized ciliopathy, and of TALS/RFMN/LWS, thus widening the clinical spectrum of *RNU4ATAC*-associated disorders and indicating ciliary dysfunction as a mechanism downstream of minor splicing defects. This finding is supported by alterations of primary cilium function in TALS and JBTS/RFMN fibroblasts, as well as by *u4atac* zebrafish model, which exhibit ciliopathy-related phenotypes and ciliary defects. Altogether, our data indicate that alteration of cilium biogenesis is part of the physiopathological mechanisms of TALS/RFMN/LWS, secondarily to defects of minor intron splicing.

## Introduction

Pathogenic variants of the *RNU4ATAC* gene, transcribed into the U4atac small non-coding RNA involved in minor splicing, are associated with three rare autosomal recessive syndromes: Taybi-Linder (TALS, also named microcephalic osteodysplastic primordial dwarfism type 1, MOPD1) (Edery *et al*, 2011; He *et al*, 2011), Roifman (RFMN) (Merico *et al*, 2015) and Lowry-Wood (LWS) (Farach *et al*, 2018) syndromes. TALS, RFMN and LWS patients are characterized by distinct patterns of skeletal dysplasia and different degrees of severity of ante- and post-natal growth retardation, microcephaly, intellectual disability, retinal dystrophy and immunodeficiency, as well as variable accompanying traits including syndrome-specific facial dysmorphism, dermatitis, hepatomegaly (RFMN), endocrine abnormalities and reduced survival (TALS) (Hagiwara *et al*, 2020; Putoux *et al*, 2016). TALS patients also display severe cerebral malformations including abnormal gyral pattern, corpus callosum abnormalities and hypoplasia of cerebellar vermis. For those who carry a specific recurrent *RNU4ATAC* mutation (n.51G>A), death usually occurs within the two first years of life, rapidly after an apparently trivial infectious episode, for yet unknown reasons.

The U4atac snRNA is a component of the minor spliceosome involved in the splicing of 935 so-called minor or U12-type introns found in 748 genes, in an up-to-date analysis of the human genome, recognized through their consensus splice site sequences (Turunen *et al*, 2013). U4atac/U6atac shares with U4/U6, despite sequence differences, a common secondary structure consisting of intra- and inter-molecular base-pairing. Some of the resulting domains such as Stem II and 5’ Stem-Loop bind essential splicing proteins. Most TALS patients carry homozygous or compound heterozygous mutations in the 5’ Stem-Loop, while all RFMN patients carry one of their two variants in Stem II, never found mutated in TALS (Benoit-Pilven *et al*, 2020). The physiopathological links between *RNU4ATAC* variants and TALS/RFMN/LWS symptoms remain largely unknown. Although it has been shown that splicing defects affect most U12-containing genes (Dinur Schejter *et al*, 2017; Heremans *et al*, 2018; Merico *et al*., 2015) to an extent that depends on the cell type (Cologne *et al*, 2019), the resulting molecular perturbations causing the developmental anomalies have not been investigated yet, due to the lack of animal models and biological hypotheses.

In the present work, we report for the first time that a homozygous pathogenic variant of *RNU4ATAC* causes a disorder with overlapping traits of Joubert syndrome (JBTS), a well-known ciliopathy, characterized by developmental delay, dysregulation of the breathing pattern, hypoplasia of the cerebellar vermis and a distinctive neuroradiologic “molar tooth sign” (MTS). This genetic finding made us consider an unsuspected link between minor splicing deficiency and cilium dysfunction, which we further confirmed in both patients’ cells and zebrafish model. Hence, *RNU4ATAC*-associated pathologies join the newly proposed group of “disorders with ciliary contribution”.

## Results

### Identification of a homozygous RNU4ATAC mutation in patients with JBTS traits

Two unrelated children suspected with having JBTS were referred to the genetic consultation. A panel targeted exome sequencing was performed, but no bi-allelic pathogenic variants or quantitative rearrangements were detected in known JBTS-related genes or other ciliary protein-encoding genes present on the ciliome/cildiag panels (see Materials and Methods). After modification of several bioinformatic parameters and the removal of a filter excluding non-coding genes, we identified a homozygous pathogenic variant, n.16G>A, in *RNU4ATAC* (GenBank: NR_023343) (Fig. 1A, Table S1). This variant was previously identified as pathogenic, either at the compound heterozygous (n=4) (Merico *et al*, 2015) or homozygous (n=2) (Dinur Schejter *et al*, 2017; Heremans *et al*, 2018) state in patients with RFMN syndrome, for whom no brain investigation was reported. Patient 1 (F1:II-2, P1) exhibited a severe developmental delay with severe axial hypotonia, microcephaly and skeletal dysplasia, associated with facial dysmorphism, retinal coloboma, nystagmus, eczema, bilateral postaxial polydactyly, genital anomalies, growth hormone deficiency, small interventricular septal defect and immunodeficiency. Brain MRI showed thin but complete corpus callosum, vermis hypoplasia, a MTS and a small posterior arachnoid cyst below the left internal vein (Table 1, Fig. 1B). Patient 2 (F2:II-3, P2), who died before two weeks of age, showed microcephaly and skeletal dysplasia, associated with an atrial septal defect, hand and feet abnormalities and ocular nystagmus (and strabismus). Brain MRI showed a short and dysgenetic vermis with enlarged retro- and infra-cerebellar cisterns, and a MTS (Table 1, Fig. 1C). Close examination of all clinical features showed that, in addition to the cerebellar and brainstem anomalies suggestive of JBTS, both patients presented with other ciliopathy traits, i.e. polydactyly and cardiac septal defects, as well as typical features of the RFMN/TALS spectrum, i.e. microcephaly, growth retardation, skeletal dysplasia and immunodeficiency (Table 1). Thus, we concluded that these patients exhibited phenotypic traits overlapping both ciliopathy and *RNU4ATAC*-disease spectra, notably RFMN, suggesting that *RNU4ATAC* loss of function secondarily results in ciliary defects.

**Figure 1.**
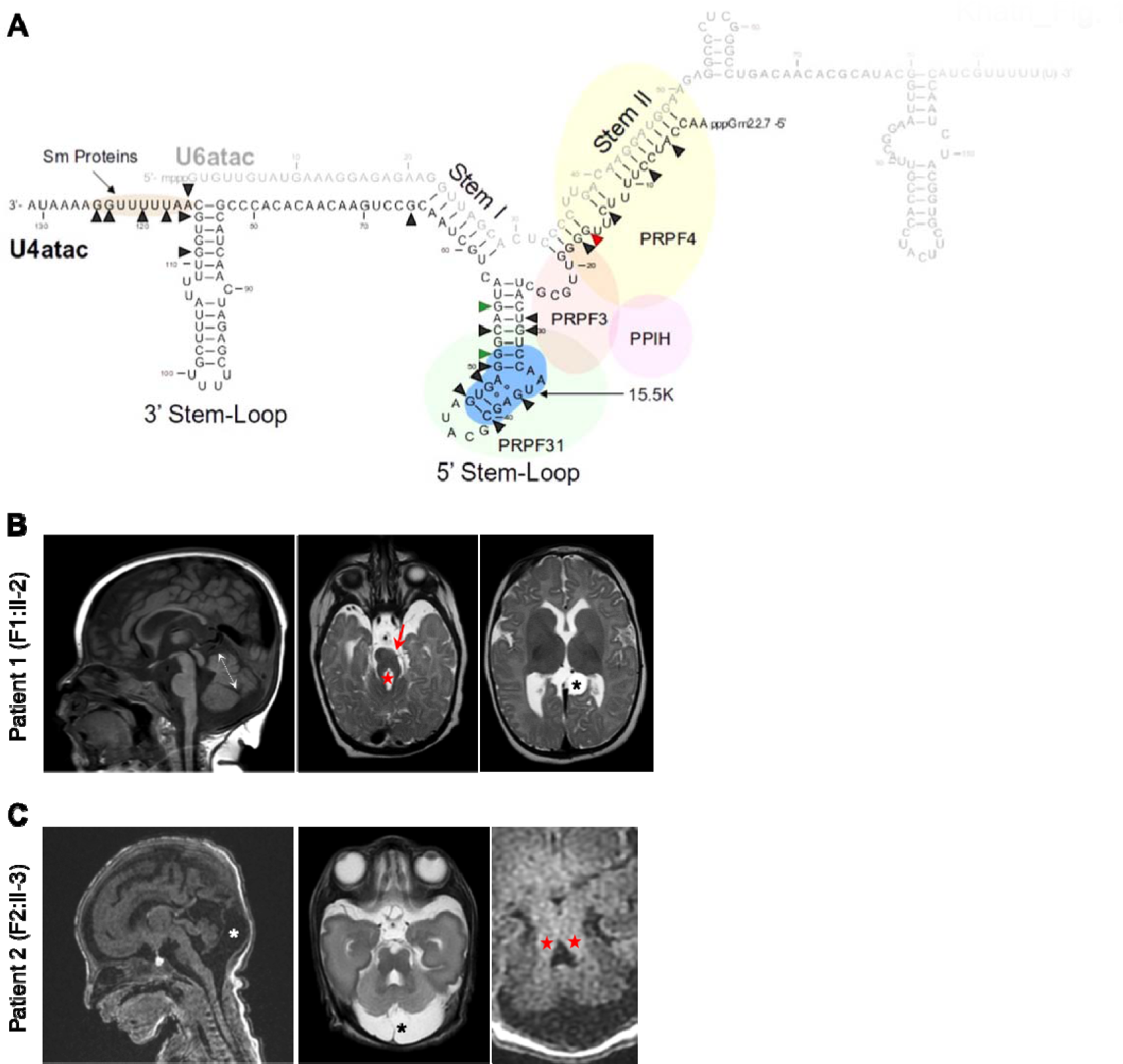
*RNU4ATAC* is mutated in two independent cases of patients with JBTS traits. **(A)** Schema of the structure of the U4atac/U6atac bi-molecule, with the main interacting proteins, showing the nucleotides mutated in patients with *RNU4ATAC*-associated syndromes. The red arrowhead points to the guanine nucleotide in position 16, which is mutated (n.16G>A) in patients with JBTS/RFMN; the two green arrowheads point to the guanine nucleotides in positions 51 and 55, the most frequently mutated nucleotides in TALS patients. **(B)** Patient 1, F1:II-2: Cerebral examination performed within first year of age. Mid-sagittal T1 weighted MR image showing a complete but short vermis suggestive of vermian hypoplasia (vermian height 25 mm, double arrow) with normal corpus callosum (left panel). Infra (middle panel) and supra (right panel) tentorial T2- weighted images demonstrating unilateral hypoplasia of the left cerebral peduncle (red arrow) as well as elongation and thickening of the superior cerebellar peduncle leading to the characteristic « molar tooth sign » (red star), and a small arachnoid cyst below the left internal cerebral vein (black asterisk). **(C)** Patient 2, F2:II-3: Cerebral examination performed in a premature baby born at few days of age. Mid-sagittal T1 weighted MR image (left panel) showing a complete but short (vermian height 12 mm, 50th centile 18,5 mm) and dysgenetic (lack of fissures) vermis with enlarged retro and infra-cerebellar cyst suggestive of Blake pouch cyst (white asterisk), well demonstrated on infra-tentorial T2-weighted image at the level of the middle peduncles (middle panel) (black asterisk). Axial T1-weighted image (right panel) showing abnormal elongated superior cerebellar peduncles (red stars) leading to typical molar tooth sign associated to abnormal fourth ventricle shape (antero-posterior diameter superior to transverse diameter) as previously reported as a prenatal feature for Joubert or Joubert-like syndrome (Quarello *et al*, 2013).

**Table 1.**
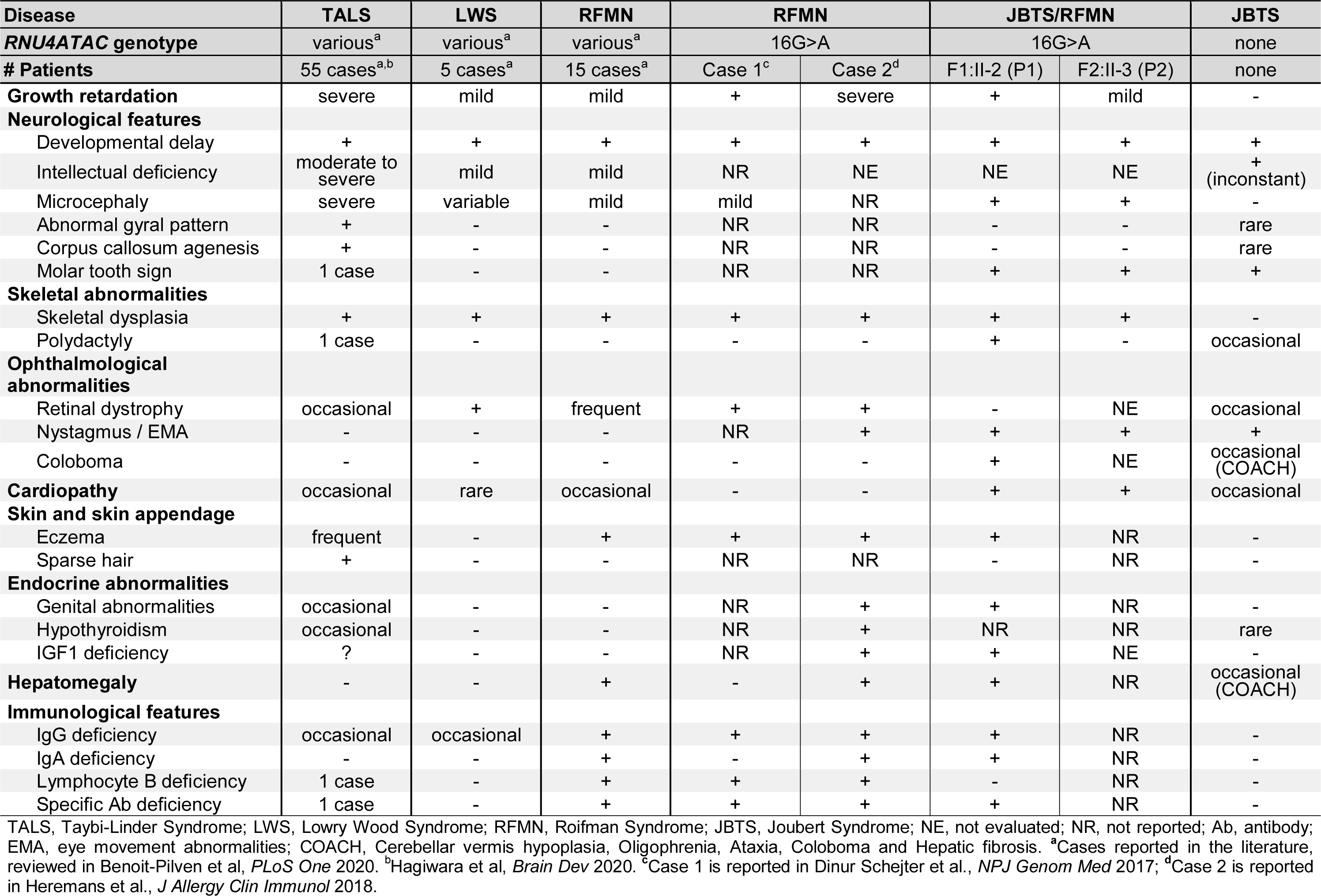
: Clinical features of patients with *RNU4ATAC* mutations

### MTS and ciliopathy traits can be found in other RNU4ATAC-mutated patients

This finding led us to conduct a thorough review of all *RNU4ATAC*-mutated published cases. A MTS had been noted in a TALS patient carrying the most frequent *RNU4ATAC* n.51G>A mutation at the homozygous state (Abdel-Salam *et al*, 2013), even though the posterior interpeduncular fossa was not as deep as in the classic MTS (G. Abdel-Salam, personal communication). Also, other ciliopathy-related traits had been observed in TALS/RFMN/LWS patients, notably retinitis pigmentosa, a cardinal feature of RFMN, and less frequently, polydactyly (TALS8 case) (Edery *et al*, 2011) and cystic kidney (TALS10 case) (Edery *et al*, 2011). Atrial or ventricular septal defects, frequently observed in TALS/RFMN/LWS patients, are also a trait commonly found in ciliopathies (Burnicka-Turek *et al*, 2016), although not as specific as other phenotypes. Therefore, the presence of ciliopathy traits in other *RNU4ATAC* patients suggests that alteration of cilium structure/function is a possible mechanism downstream any mutations of *RNU4ATAC*, and is not restricted to n.16G>A pathogenic variant.

### Cilium-related genes are over-represented among human U12 genes

To better apprehend the link between minor splicing and cilium function, we conducted a gene ontology (GO) enrichment analysis of U12 genes. Relatively few studies addressed the functions of U12 genes in recent years, since their full identification has been facilitated both by the completion of the human genome sequence and the development of computational predictive tools. To the best of our knowledge, only three GO enrichment analyses have been performed, one based on functional gene-sets and mouse phenotypes from the Mammalian Phenotype Ontology resource (Merico *et al*, 2015), one on the most differentially expressed U12 genes in adult human tissues (Olthof *et al*, 2019) and the last on a subset of U12 genes associated with a disease (Olthof *et al*, 2020). Here, we used a different approach and performed a GO term enrichment analysis of the U12 genes *versus* the U2 genes, using the entire set of genes with GO annotations (n=618 and 19,939 for U12 and U2 genes, respectively). An enrichment is observed if more than 3% (618/20,557) of U12 genes are present in a GO term. Hence, we found 243 enriched biological processes and 96 enriched cellular components with a p-value ≤0.05 (Fig. 2A, Tables S2, S3). The most enriched biological process is the *membrane depolarization during action potential* (p-value = 5.2e-15, with 40% of U12 genes), which partially overlaps the other GO terms *neuronal action potential* (p=5.5e-13, 40%), *regulation of ion transmembrane transport* (p=1.2e-12, 8%), *calcium ion transmembrane transport* (p=4.3e-6, 9%) and *sodium ion transmembrane transport* (p=5.2e-6, 10%). Interestingly, the *non-motile cilium assembly* process (p=6.8e-6, 18%) was the thirteenth most significant term (red in Fig. 2A, Table S2). Other significant terms explicitly related to cilia were identified, namely *intraciliary transport involved in cilium assembly* (p=1.2e-3, 15%), *ciliary basal body-plasma membrane docking* (p=1.6e-2, 7%) and *protein localization to ciliary transition zone* (p=2.8e-2, 22%). Similarly, GO term enrichment analysis of cellular components revealed an unexpected amount of U12 gene-encoding proteins that localize at the basal body (p=1.4e-5, 11%), the ciliary base (p=4.3e-3, 14%) and the ciliary tip (p=1.0e-2, 11%) (Table S3). Of note, the opposite comparison of U2 *vs* U12 genes did not reveal any cilium-related terms (Table S4).

**Figure 2.**
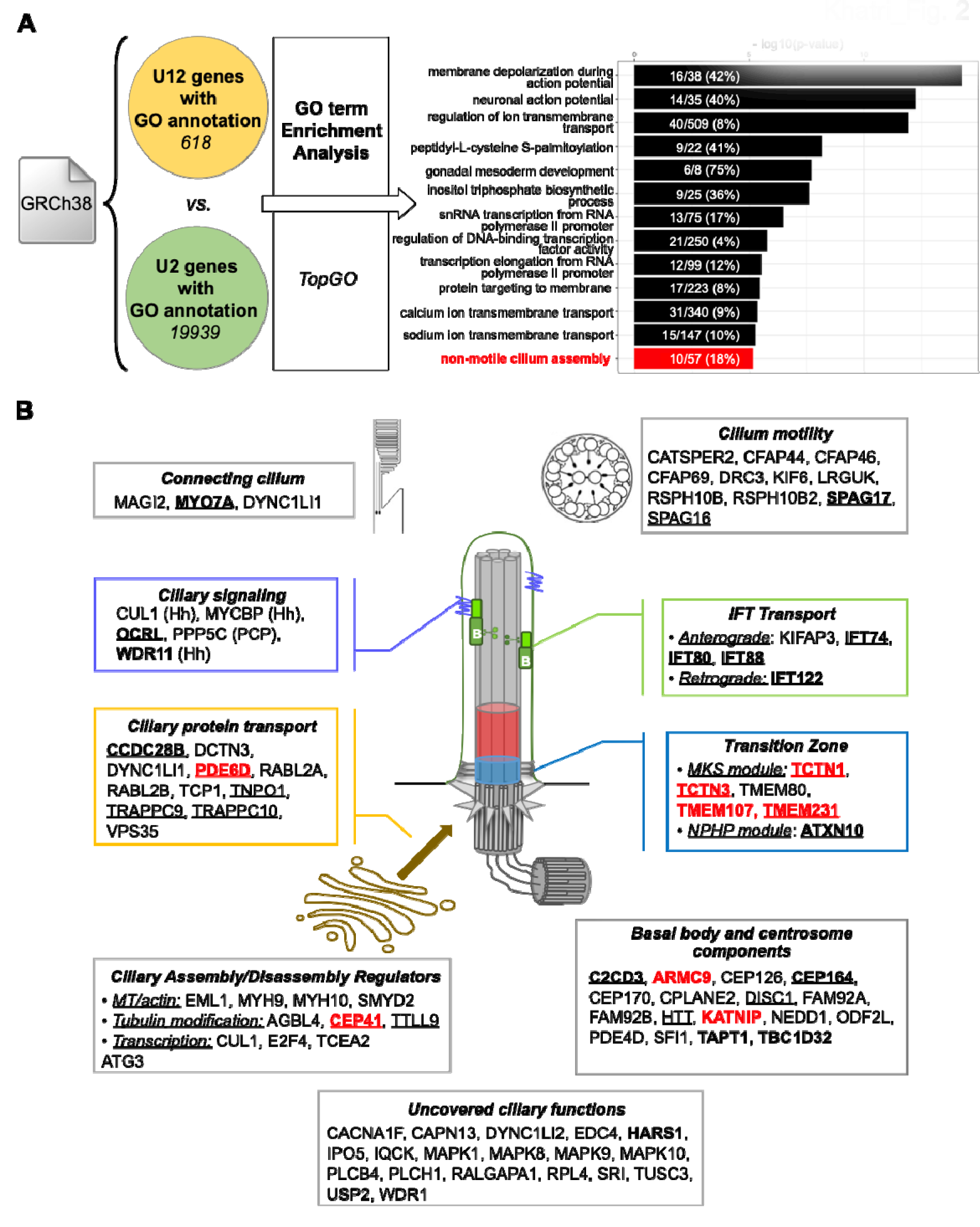
The non-motile cilium assembly pathway is enriched among U12 genes. **(A)** GO term enrichment among U12 intron-containing genes. Comparison of 618 U12 and 19,939 U2 genes with Gene Ontology annotations, using the TopGO R package, allowed to identify 80 enriched biological processes with a p-value<0.05, of which the 13 most significant are shown. Numbers in bars indicate the ratio of U12 gene to the total gene number belonging to each GO term; in brackets the percentage of U12 genes among the GO term. **(B)** Schema representing the localisation and/or function of the 86 cilium-related U12 genes. Underlined are the “Gold standard” genes (van Dam *et al*, 2013), in bold the genes associated with a ciliopathy, and in red those associated with JBTS. Hh, hedgehog pathway; PCP, Planar Cell Polarity pathway.

We then examined the broad class of cilium-related genes, bearing in mind that 2.0% of the genes are of the U12-type category in the human genome (748 U12 vs 37,173 U2 genes). Since we observed 7.6% (23 U12 genes vs 279 U2 genes, Table S5) of U12 genes among the curated ciliary “Gold_Standard” genes (van Dam *et al*, 2013), we concluded that there is an enrichment of U12 genes among them (hypergeometric test p-value = 1.0e-8). By broadening this list through the compilation of several databases (1426 genes in total, see Materials and Methods), we identified 86 U12 genes (86/1426, 6%) within this in-house ciliome database (Fig. 2B, Table S5). Finally, among the 329 ciliopathy-related genes (Lovera & Lüders, 2021; Reiter & Leroux, 2017; Wheway *et al*, 2019) (Table S5), 23 (∼7%) are U12 genes (hypergeometric test p-value = 5.2e-8). Strikingly, nearly a third of them are associated with JBTS (*KIAA0556, CEP41, TCTN3, ARMC9, PDE6D, TMEM107, TCTN1, TMEM231*) (Fig. 2B, Table S5), corresponding to 21.6% (8/37) of all known JBTS-associated genes.

Altogether, these enrichment analyses indicate that U12-intron mis-splicing is likely to impair the expression of many genes crucial for cilium formation and/or function, thus suggesting a possible explanation for the ciliary contribution in *RNU4ATAC*-associated disorders.

### JBTS/RFMN RNU4ATAC mutation results in U12 intron retentions

To validate the presence of intron retention in JBTS/RFMN patient cells, we conducted a transcriptomic study and qRT-PCR analyses. For this, we followed the approach we used in our recent in-depth analysis of transcriptomes of three different cell types (fibroblasts, amniocytes, lymphocytes) from seven TALS patients, which showed a specific and global alteration of minor splicing (U12 intron retentions (IR) mostly) while gene expression profiles were unchanged (Cologne *et al*, 2019). Hence, the RNA-seq experiment carried out on fibroblasts derived from the skin biopsy of patient P1 revealed, like previously in TALS samples, about the same amount of U12 expressed genes (∼450), of which a small proportion (10%) showed differences of percent-spliced-in (PSI) values of the U12 intron greater than 10% compared to the control (Fig. 3A, Table S6). These U12 intron retentions were not associated to drastic alterations of gene expression level (Supplemental Table S6). Although biological replicates (i.e. fibroblasts issued from other patients with n.16G>A mutation) would be needed to draw conclusions, the levels of U12 intron retentions, globally weak (maximum deltaPSI of ∼30%), are reminiscent of those seen in TALS samples, with most U12 genes (∼60%) having a dPSI<10% (Cologne *et al*, 2019). We further performed qRT-PCR analyses to specifically analyse the U12 intron retention of some ciliary genes (*RABL2A, TMEM107, TMEM231, TCTN1, IFT80*, see Fig. 2B) and observed, for all of them, a higher intron retention level in patients’ cells than in controls, with a stronger effect seen in P1 compared to a TALS patient cells (Fig. 3B). It is noteworthy that qRT-PCR analyses evidence stronger IR than RNA-seq, particularly for *RABL2A* and *TMEM107*, which may be explained by the low coverage of their U12 intron boundaries by NGS.

**Figure 3.**
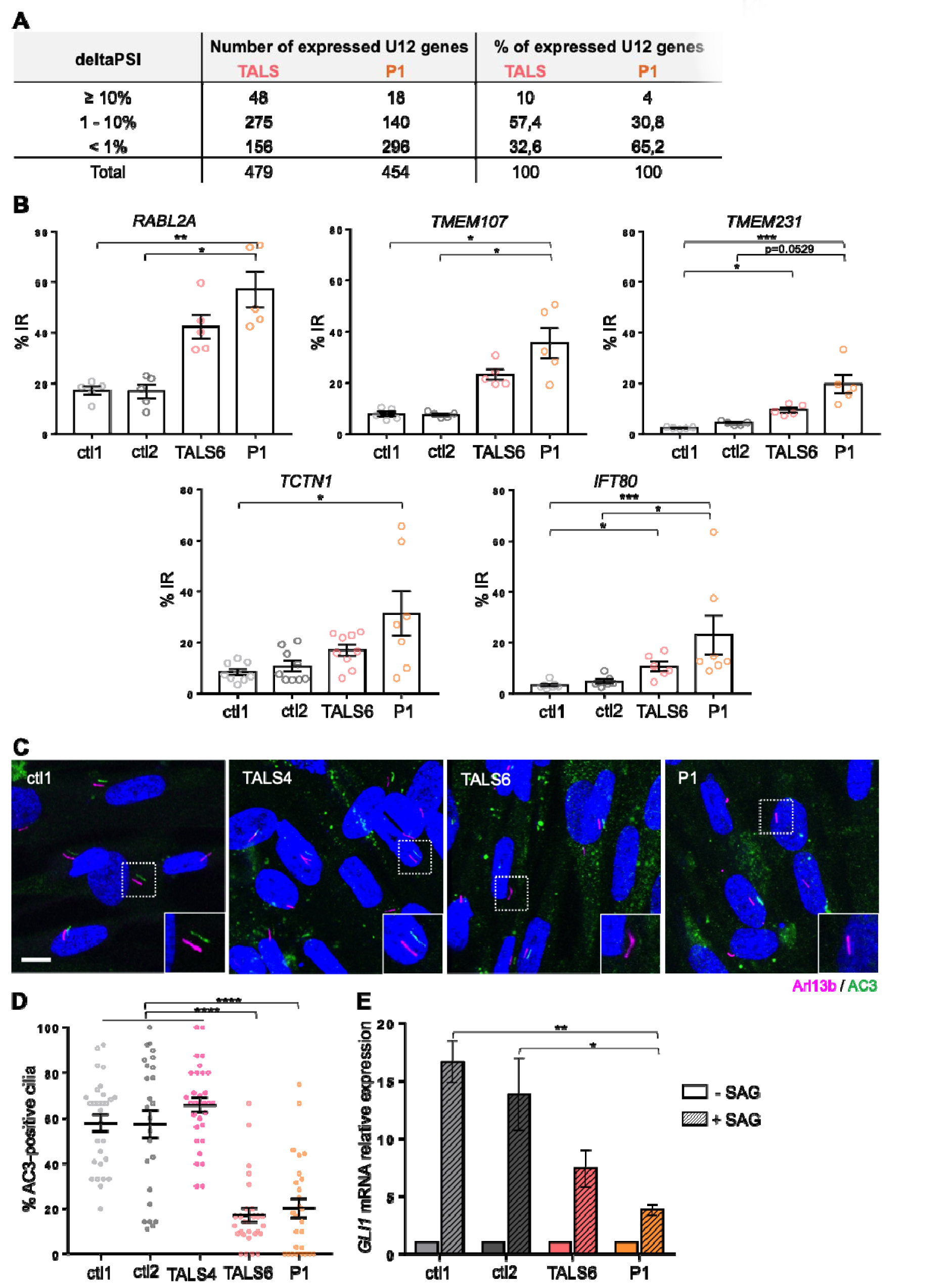
TALS and JBTS/RFMN-related *RNU4ATAC* mutations lead to U12 intron retention in ciliary genes and result in ciliary function alteration in patient’s fibroblasts. **(A)** Table summarising the results of the analysis of U12 intron retention in the transcriptomic data of Patient 1 (P1) *vs* ctl1 fibroblasts, in the same format as those we previously published (Cologne *et al*, 2019). Indicated are the number and percentage of U12 genes presenting differences of percent-spliced-in (PSI) values (deltaPSI) greater than 10%, between 10 and 1%, or below 1% between patient and control cells. **(B)** qRT-PCR analyses of inton retention (IR) of U12 ciliary genes in control, TALS6 and P1 fibroblasts. Graphs show the mean ± s.e.m. of at least 3 independent experiments. *p<0.05, **p<0.01, ***p<0,001, using Kruskal-Wallis test with Dunn’s multiple comparisons. **(C)** Confocal images of age- and sex-matched control, TALS and P1 patients’ fibroblasts derived from skin biopsy. Adenylate cyclase (AC3, green) is co-labelled with Arl13b (cilium marker, magenta). Scale bar, 10 µm. **(D)** Quantification of the percentage of AC3- positive cilia seen in (C). Graphs show the mean ± s.e.m. of at least 6 fields of 5 independent experiments (total number of analysed cells >200). ****p<0.0001, using Kruskal-Wallis test with Dunn’s multiple comparisons. **(E)** qRT-PCR analysis of the expression of the hedgehog target gene *GLI1* upon SAG treatment (+ SAG). Graphs show the mean ± s.e.m. of 4 to 5 independent experiments. **p<0.01, *p=0,0499, using Kruskal-Wallis test with Dunn’s multiple comparisons.

Altogether, these results confirmed that the n.16G>A mutation, similarly to other *RNU4ATAC* mutations associated with TALS, causes U12 intron retention in patient fibroblasts, including in U12 ciliary genes.

### Primary cilium function is altered in RNU4ATAC-associated JBTS/RFMN and TALS fibroblasts

To investigate whether intron retention had an impact on centrosome structure and ciliogenesis, we studied patients’ fibroblasts, a well described and favored ciliated cell model, issued from JBTS/RFMN patient P1 and from two TALS patients carrying the n.51G>A homozygous mutation (patients TALS4 and TALS6 described in Edery *et al*, 2011). Analysis of the centrosome structure by ultrastructure expansion microscopy revealed no alteration of centriole composition; however, a slight increase of centriole length was observed in P1 fibroblasts (Fig. S1A-B). Regarding ciliogenesis, we analysed the percentage of ciliated cells and cilium length, based on immunostainings of cilia with an antibody anti-Arl13b, which revealed slight ciliogenesis alterations in the two TALS cell lines, but not in P1 patient cells (Fig. S1C-E). We then evaluated cilium function and explored the ciliary localisation of the protein adenylate cyclase III (AC3), which plays a crucial role in signal transduction of G-protein coupled receptors (GPCR) through the production of the second messenger cAMP. We observed a dramatic loss of AC3 into the cilium in TALS6 and P1 fibroblasts compared to control and TALS4 cell lines (Fig. 3C, D). We also explored hedgehog (Hh) signalling, which is frequently altered in JBTS models. For this, we analysed the expression of the target gene *GLI1* upon activation of the pathway with the smoothened agonist SAG. As compared to control cell lines, *GLI1* expression was twice less activated upon SAG treatment in both TALS6 and P1 fibroblasts (Fig. 3E). Hence, altogether these results indicate that loss of U4atac function impairs primary cilium function (AC3-dependant cAMP signalling, Hh) rather than ciliogenesis. This could be explained by the fact that six U12 genes encode components of the transition zone, an essential filter barrier that controls entry and exit of ciliary proteins in and out of the cilium. Also, it is noteworthy that primary cilium alterations are variable, more or less subtle from one to another individual, which can result from the global low level of U12 intron retentions seen in fibroblasts, indicating that this cell type may not be the most suitable for such analyses.

### Deficiency of u4atac in zebrafish leads to ciliary defects

To evaluate the importance of minor splicing during development, we turned to the zebrafish, a widely used animal model to study genetic diseases, including ciliopathies (Song *et al*, 2016). Its genome includes 710 U12 introns present in 655 genes in the latest genome assembly (GRCz11), as revealed by a computational scan we performed using U12 intron scoring matrices (Alioto, 2007). These figures are in the same range as those found in humans, and most of the U12 zebrafish genes (544/655) are orthologs of human genes. Zebrafish has two orthologs for *RNU4ATAC*, and by combining the analysis of sequence and secondary structure conservation, expression throughout development and phenotype of CRISPR/Cas9-mediated knock-out lines (see Supplemental Methods, Fig. S2, S3), we concluded that *rnu4atac* on chromosome 11 was the sole functional ortholog of human *RNU4ATAC*.

Pathogenic variants in humans are hypomorphic and full deletion of rnu4atac_chr11 in zebrafish precluded the analysis of phenotypes beyond 22 hpf due to growth arrest and lethality. We therefore opted for a morpholino-based approach and designed two morpholino oligonucleotides (MO) that targeted two distinct functional domains of u4atac, the 5’ Stem Loop and Stem II (Fig. S4A). Both MO led to a wide range of developmental anomalies whose severity was dose-dependent and concomitant with an increasing level of U12 intron splicing deficiency (Fig. 4A-G, Fig. S4B-E). Among the most penetrant phenotypes (>50% of MO-injected embryos), we observed body axis curvature, pronephric cysts and otolith defects as well as cardiac dysfunction (Supplemental Methods, Movies S1, S2) and absence of touch response. To a lesser extent (<30%), u4atac morphants also exhibited microcephaly and brain hemorrhages (Fig. 4A-G, Fig. S4D). All these phenotypes were rescued by the co-injection of human WT U4atac snRNA (Fig. 4G, Fig. S4D), in accordance with the highly similar bi-dimensional predictions of human U4atac/U6atac, zebrafish u4atac/u6atac and human U4atac/zebrafish u6atac duplexes (Fig. S2D). Strikingly, the three most penetrant phenotypes (i.e. body axis curvature, pronephric cysts and otolith defects) are hallmarks of cilium dysfunction in zebrafish, thus strengthening the link between u4atac, minor splicing and cilium function. For further evidence, we analysed cilium structure in the central canal, pronephros and otic vesicle. Cilium dysfunction in the central canal results in cerebrospinal fluid flow alteration and Reissner’s fiber mis-aggregation, shown to be the origin of the ventral body curvature phenotype (Cantaut-Belarif *et al*, 2018). In the pronephros, motile cilia, notably those of multiciliated cells, allow fluid flow in renal tubules; their impairment leads to fluid accumulation, tubule luminal expansion and cyst formation in glomeruli (Kramer-Zucker *et al*, 2005). In the otic vesicle, motile cilia participate in otolith formation, favoring the binding of otolith precursor particles to the tip of kinocilia (Wu *et al*, 2011). We analysed motile cilia in the central canal by performing confocal live imaging of *Tg(bactin:arl13bGFP)* transgenic line. We observed a drastic decrease of cilium number in morphants (Fig. 4H), associated with defective cilium beating (Fig. 4H, I). In pronephros, we observed tubule dilations, mostly of the proximal straight tubule, that were accompanied by a lack of multiciliated tufts (Fig. 4J). Similarly, the number of cilia lining the otic vesicle was decreased (Fig. 4K, L). Finally, analysis of cilium formation at the nasal pit, at 48 hpf, revealed a significant alteration of the ciliary tuft in u4atac morphants compared to control embryos (Fig. 4M). All these ciliary defects were partially or totally rescued by the expression of human WT U4atac snRNA (Fig. 4G-M).

**Figure 4.**
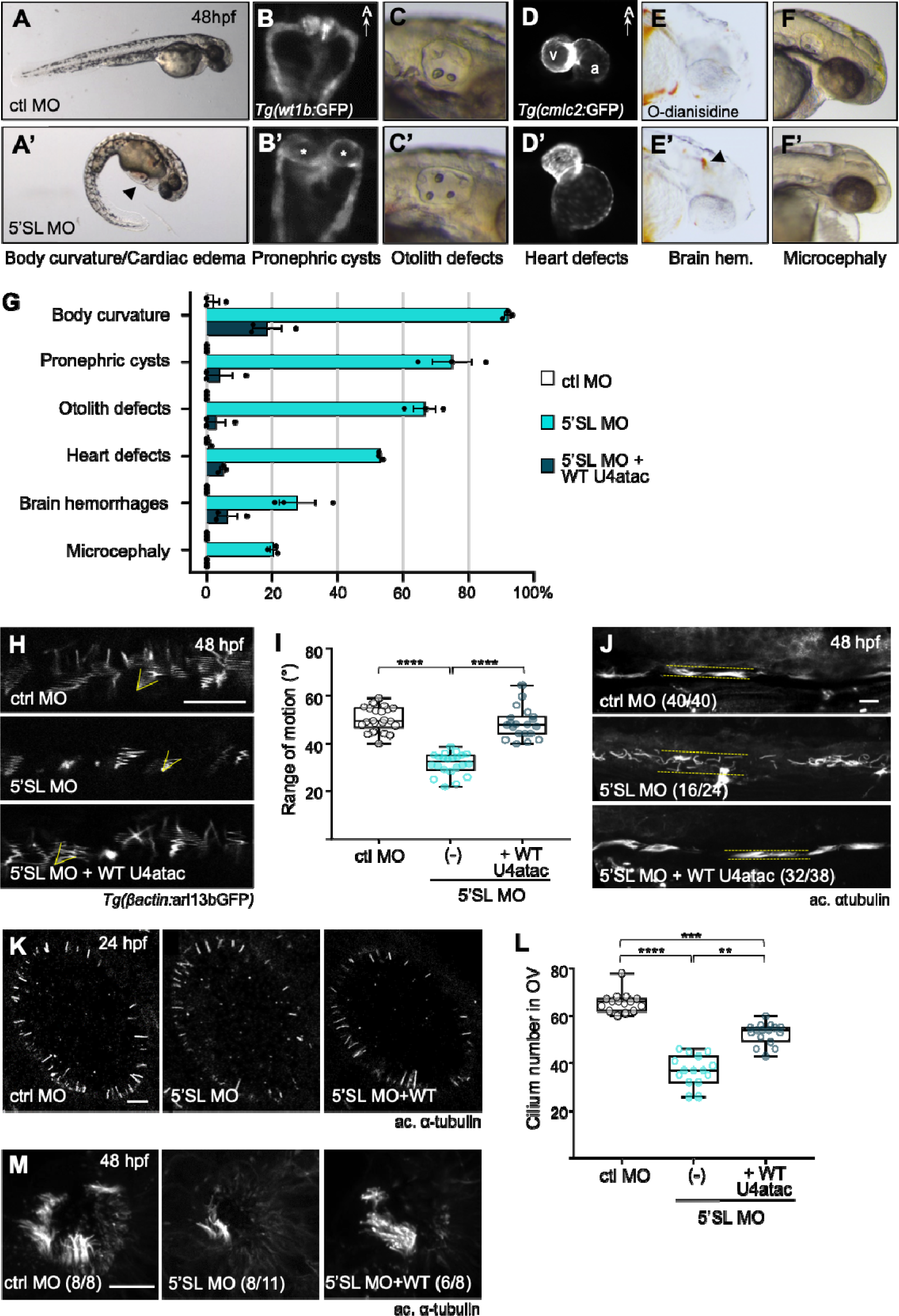
Deficiency of u4atac leads to cilium alterations in zebrafish. **(A-F)** Global morphology of control (ctl) (upper panel) and u4atac 5’SL (lower panel) morpholino (MO)-injected embryos at 48 hpf. u4atac morphants are characterized by body curvature and cardiac edema (A’, arrowhead); cysts (B’, asterisks) in *Tg(wt1b:GFP)*-labelled glomeruli; anomalies of otolith morphology/number in otic vesicle (C’); alterations of cardiac morphology with atrial hypertrophy, as seen with *Tg(cmlc2:GFP)* transgene (D’); blood hemorrhages in brain, as visualised by O-dianisidine staining (E’, arrowhead); and microcephaly. Dorsal views in B-B’ and ventral views in D-D’, with anterior to the top. **(G)** Percentage of embryos displaying each of the phenotypes shown in A-F, following injection of control MO, u4atac 5’SL MO alone or with human WT U4atac snRNA. Graph shows the mean ± s.e.m. of three independent experiments (total number of embryos : >300). p<0.01, t-tests comparing 5’SL MO to ctl MO, and 5’SL MO + WT to 5’SL MO. **(H)** Live confocal imaging of motile cilia in central canal of MO-injected *Tg(bactin:arl13bGFP)* transgenic embryos, at 48 hpf. Slow acquisition speed of motile cilia produces imaging artefacts, delineating the range of motion of each cilium (yellow angles). Images show the maximum intensity projection of z-stack. **(I)** Quantification of the angle defined by the range of motion of motile cilia in control MO (n=22), u4atac 5’SL MO alone (n=22) or with human WT U4atac snRNA (n=20) injected embryos, as described in (H). Box-and-whisker plots show in the box the median and the 25th-75th percentiles, and in whiskers the minimum to the maximum values. Values are the mean ranges of motion of each single embryo analysed in two independent experiments. ****p<0.0001, by Kruskal-Wallis test with Dunn’s multiple comparisons test. **(J)** Immunostaining of cilia (acetylated α-tubulin) in the proximal straight tubule of control MO, u4atac 5’SL MO alone or with human WT U4atac snRNA injected embryos, at 48 hpf. In brackets, the number of embryos exhibiting the phenotype shown in the image over the total amount of analysed embryos. Dotted lines delineate the tubule lumen. Images show the maximum intensity projection of z-stack. **(K)** Immunostaining of cilia (acetylated α-tubulin) in the otic vesicle of control MO, u4atac 5’SL MO alone or with human WT U4atac snRNA injected embryos, at 24 hpf. Images are single optical slice selected in the middle of z-stack. **(L)** Quantification of the cilium number in otic vesicle of control MO (n=15), u4atac 5’SL MO alone (n=15) or with human WT U4atac snRNA (n=16) injected embryos, as described in (K). Box-and-whisker plots show in the box the median and the 25th-75th percentiles, and in whiskers the minimum to the maximum values, which were obtained in two independent experiments. ****p<0.0001, ***p<0.005, **p<0.01 by Kruskal-Wallis test with Dunn’s multiple comparisons test. **(M)** Immunostaining of cilia (acetylated α-tubulin) at the nasal pit of control MO, u4atac 5’SL MO alone or with human WT U4atac snRNA injected embryos, at 48 hpf. In brackets, the number of embryos exhibiting the phenotype shown in the image over the total amount of analysed embryos. Images show the maximum intensity projection of z-stack. Scale bars, 10 µm. a, atrium; v, ventricle; hem, hemorrhages.

Altogether, these data indicate that loss of function of u4atac is associated *in vivo* with ciliary defects, which are likely secondary to minor splicing alterations.

### JBTS/RFMN and TALS U4atac mutations lead to ciliopathy-related phenotypes in zebrafish

To have a better assessment of the impact of human U4atac variants on embryonic development processes linked to cilium function, we took advantage of the zebrafish model and conducted rescue experiments. We co-injected u4atac 5’SL MO with human U4atac carrying either the n.16G>A, n.51G>A or n.55G>A mutation. These mutations have been identified at the homozygous state in 4, 31 and 8 patients or fetuses respectively (Benoit-Pilven *et al*, 2020). They are associated with slightly different presentations (Table S7) and large differences in life expectancies: mean age at death is 10 months for patients homozygous for n.51G>A (three surviving patients, ≤ 2 years old), while 3/4 and 4/8 of those with n.16G>A or n.55G>A were still alive at the time of publication, their age varying from 2 to 40 and 2 to 20 years old, respectively.

Stability of injected human snRNA was validated by Northern blot (Fig. S4F), and the above-mentioned phenotypes were recorded. Regarding the body curvature, we observed that 16A, 51A and 55A snRNAs had little capacity to rescue the phenotype, with nevertheless a gradation between their effect, 55A snRNA co-injected-embryos having a slightly milder phenotype than those co-injected with 16A, themselves a little better than those co-injected with 51A snRNAs (Fig. 5A, B). Considering all other phenotypic features, the 51A snRNA seemed to have the lowest ability to rescue the anomalies compared to 16A and 55A, the latest corresponding to the less severe mutation (Fig. 5C). We also explored the impact of the mutations at the molecular level by analysing in zebrafish the U12 intron retention of six cilium-related genes (*rabl2, tmem107l, tmem231, ift80, tctn1, pde6d*), among which the five genes tested in TALS and P1 patients’ cells. For all of them, the U12 intron was strongly retained in 5’SL MO-injected embryos, and mis-splicing could be rescued to the level of control by the co-injection of WT U4atac. Consistent with the observed phenotype, n.51G>A had a more severe impact on minor splicing than n.16G>A, and then n.55G>A (Fig. 5D).

**Figure 5.**
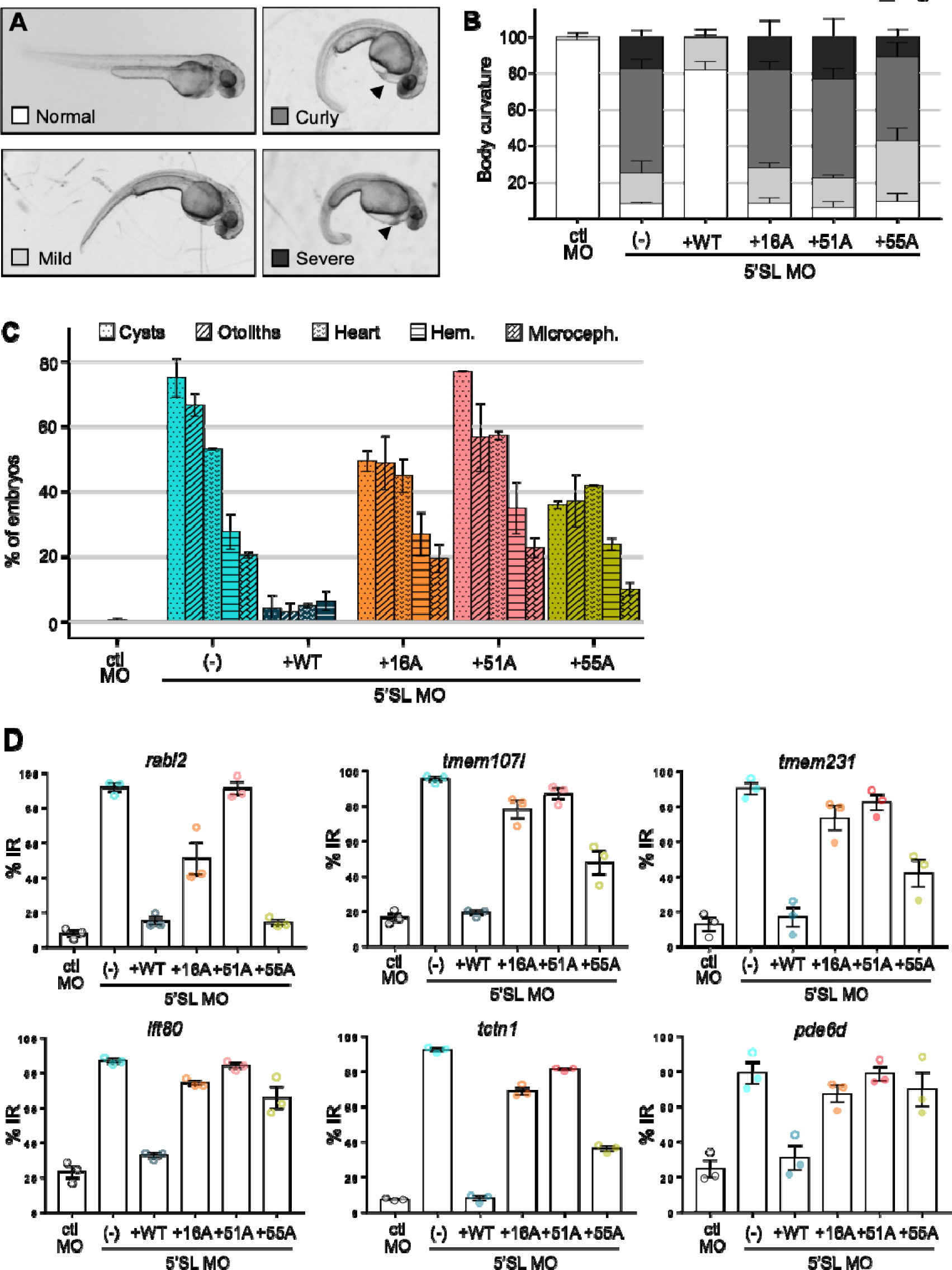
Co-injected JBTS/RFMN and TALS U4atac mutants lead to ciliopathy-related phenotypes in zebrafish. **(A)** Global morphology of embryos co-injected with 5’SL MO and JBTS/RFMN (16A) or TALS (51A, 55A) -related U4atac variants. A varying severity of body curvature could be observed (mild, curly, severe). **(B)** Quantification of embryos exhibiting the various severities of body curvature shown in (A). **(C)** Percentage of embryos displaying each of the phenotypes shown in Fig. 4A-F, following co-injection of 5’SL MO with human WT or mutated U4atac snRNA. Graph shows the mean ± s.e.m. of three independent experiments (total number of embryos: 180). **(D)** qRT-PCR analysis of U12 intron retention (IR) in six cilium-related genes in embryos co-injected with 5’SL MO and human WT or mutated U4atac snRNA. Graphs show the mean ± s.e.m. of three independent experiments.

In conclusion, JBTS/RFMN and TALS *RNU4ATAC* pathogenic variants all lead to ciliary defects in zebrafish, due to a deficiency of minor splicing. Of note, we observed a gradation effect of the variants, sustaining a phenotype/genotype correlation that will need to be further explored.

## Discussion

We report here for the first time the identification, in two independent cases suspected of having JBTS, of a homozygous pathogenic variant of *RNU4ATAC*, a gene previously involved in three rare syndromes (TALS, RFMN, LWS) characterized by microcephaly, short stature and other specific features. In line with this finding, we found that cilium-related genes are over-represented among human U12 genes, that several ciliary genes are impacted by defective minor splicing and that primary cilium function is altered in TALS and JBTS/RFMN fibroblasts. Furthermore, the zebrafish *u4atac* models present with ciliopathy-related phenotypes and ciliary defects that could be rescued by WT but not by U4atac carrying TALS and JBTS/RFMN variants.

Quite remarkably, the two infants we describe here have a complex syndromic disorder combining both traits seen in JBTS patients (MTS, hypotonia, nystagmus/strabismus, septal defects, polydactyly/clinosyndactyly) and in TALS/RFMN/LWS patients (microcephaly, skeletal dysplasia, growth retardation, immunodeficiency, eczema). They harbour a homozygous *RNU4ATAC* pathogenic variant, n.16G>A, the only one previously found at the homozygous state in Stem II in two typical RFMN patients (a six year old girl (Dinur Schejter *et al*, 2017) and the 38 year old male (Heremans *et al*, 2018)), while all others harboured compound heterozygous variants, one located in Stem II and the other elsewhere in U4atac. To explain the phenotypic variability among patients homozygous for n.16G>A, one could consider that other genetic alterations are responsible for the ciliopathy-related traits seen in the JBTS/RFMN patients reported here, even if all of them were found at the heterozygous state (Table S1). Nevertheless, it is noteworthy that one of the two published n.16G>A homozygous RFMN patients presented with “eye motility problems” (Table 1) (Heremans *et al*, 2018), a feature previously unseen in *RNU4ATAC*-associated pathologies but common in JBTS. As for the MTS, most RFMN patients present with a mild microcephaly and borderline-to-mild intellectual impairment, so that brain MRI is not systematically performed. Based on our findings, we suggest that the two RFMN previously published patients should have a thorough investigation of their brain morphology to explore the possible presence of a MTS. In addition, as ciliopathy-related traits have been observed in some TALS/RFMN/LWS patients (including a MTS in one of them), the overlap with ciliopathies is clearly not restricted to *RNU4ATAC*-associated JBTS cases, even if it might be greater for n.16G>A homozygotes or more largely for carriers of two pathogenic variants in U4atac Stem II. In agreement with this, a recent disease scoring system based on the occurrence of ciliopathy-like phenotypes produced high scores for RFMN and TALS (respectively 267^th^ and 428^th^ highest score among 6058 analysed disorders), making them likely candidates for disorders with ciliary contribution (Lovera & Lüders, 2021). Moreover, we bring up the identification of the homozygous c.2953A>G variant in the *RTTN* gene, encoding the centrosome/cilium protein rotatin, in a child presenting with TALS features but *RNU4ATAC*-negative (Guguin *et al*, manuscript in preparation). Our finding correlates with another published case (Grandone *et al*, 2016), and strongly supports the notion that many of the developmental abnormalities seen in TALS patients could be due to cilium/centrosome dysfunction.

Additional evidence is provided by the zebrafish *u4atac* model. As seen in patients, MO-mediated deficiency of u4atac led to phenotypes that recapitulate features both associated with TALS/RFMN/LWS (microcephaly, brain hemorrhages (Abdel-Salam et al, 2013, 2011), cardiac defects), and with ciliopathies (body axis curvature, pronephric cysts and otolith defects). Brain hemorrhages could even fall in this last category as they have been shown to result from ciliary defects of endothelial cells in zebrafish (Eisa-Beygi et al, 2018; Kallakuri et al, 2015). Our results demonstrate that the ciliopathy-related phenotypes are indeed due to ciliary defects in the central canal, pronephros, otic vesicle and olfactive bulb. These phenotypes could be mostly rescued by co-injection of the human WT U4atac snRNA but not, or to a lesser extent, by snRNA molecules carrying JBTS/RFMN or TALS-associated mutations. These results thus strongly support the fact that several types of pathogenic variants of *RNU4ATAC* can result in ciliopathy-related phenotypes.

By identifying a homozygous *RNU4ATAC* mutation in two patients with emblematic ciliopathy traits, we shed light on a new “second-order” mechanism leading to ciliary defects, i.e. splicing of minor introns. Second-order ciliopathies are caused by mutations in genes that have an indirect role in cilium formation or function (Reiter & Leroux, 2017), among which some are involved in different aspects of RNA metabolism. As examples, we can cite *DDX59* encoding a RNA helicase or *BICC1* coding for a RNA-binding protein involved in Oro-Facial-Digital (OFD) syndrome and cystic renal dysplasia, respectively (Shamseldin et al, 2013; Kraus et al, 2012). More recently, a subunit of the snRNA-processing complex Integrator, INTS13, has been linked to a complex developmental ciliopathy (Mascibroda *et al*, 2020) while other subunits were shown *in vitro* to be required for ciliogenesis (Jodoin et al, 2013). In all the second-order ciliopathies linked to the deficiency of an RNA-processing function, including minor splicing unravelled here, it remains to be explained why ciliary genes are specifically affected and how they drive the phenotype. In the case of minor splicing, the high proportion of U12 genes encoding ciliary components (at least 86, i.e. 12% of the total number of U12 genes) is likely to be an important factor, combined to the fact that the cilium is a complex sensory machine involved in transducing a wide range of extracellular stimuli into cellular responses essential for organogenesis. Further investigations are needed to address these questions, taking into account that defective minor splicing results in much more complex outcomes than the simply decreased expression of genes with U12 intron retention (due to transcript degradation by nuclear retention or non-sense mediated decay). A recent review pointed out the crosstalk mechanisms between major and minor spliceosomes, that may vary depending on cell type and throughout development (Akinyi & Frilander, 2021). Global transcriptomic analyses of splicing events are thus definitely needed to understand the impact of *RNU4ATAC* mutations, and these studies will be carried out in the zebrafish model, as well as in human development models derived from induced pluripotent cells.

To conclude, our work is of significant impact for both JBTS and TALS/RFMN/LWS patients, notably regarding the genetic diagnosis and counselling. Due to its non-coding status, *RNU4ATAC* is still under-considered among the clinically relevant genes. Not only should it be evaluated in the JBTS cases lacking a molecular diagnosis, but given the pleiotropy of the clinical signs associated with this gene, one should remain open when assessing its potential pathogenic variants in patients with rare diseases. Also, our work widens the fields of research on the physiopathological mechanisms underlying the *RNU4ATAC*-associated diseases.

## Materials and Methods

### Targeted next-generation sequencing

Sequencing of genes involved in ciliary diseases or cilia function was performed using a custom SureSelect capture kits (Ciliome V3c (1339 genes) (Failler *et al*, 2014; Thomas *et al*, 2012) or CilDiag (170 genes)) (Agilent Technologies, Santa Clara, CA, USA). Libraries were then sequenced using a HiSeq or NextSeq sequencers (Illumina, San Diego, CA, USA), respectively. Resulting paired-end reads were mapped on the GRCh37 reference genome using Burrows-Wheeler Aligner (Illumina). Bioinformatic analysis was performed using the Genome Analysis Toolkit, SAMTools and Picard Tools, in accordance to best practices issued by the Broad Institute. Variants were annotated using a software developed by the Paris Descartes University Bioinformatics platform. As a first screen, only nonsense, frameshift, splice site and missense variants, leading to an abnormal protein sequence, were retained; variants in non-exonic regions, pseudogenes, UTRs or with a frequency above 1% in public databases (dbSNP, 1000 genomes project, ExAC), or frequent in an in-house database of previously sequenced patients, were excluded. The functional consequence of missense variants was predicted using in silico tools, namely PolyPhen-2, SIFT, Mutation Taster, and CADD. Since this first screen turned out to be negative, i.e. identification of bi-allelic variants in one gene, variants in non-coding genes were thus analysed. Of note, panel analysis did not reveal any possible quantitative rearrangements, either it be gain or loss, in JBTS genes present in each of the panels. Variants identified by NGS were then confirmed by Sanger sequencing.

### Bioinformatic GO term analyses

All the sequences were analyzed using the Ensembl Genome Browser. Nucleotide and amino acid sequences were compared to the non-redundant sequences present at the NCBI (National Center for Biotechnology Information) using the Basic Local Alignment Search Tool (BLAST). Secondary structure prediction for snRNA duplexes was performed by using RNAstructure (Reuter & Mathews, 2010).

U12 genes were identified by running the U12db (Alioto, 2007) annotation pipeline on the GRCh38 version of the human genome, with the gencode38 version of the annotations, or the GRCz11 version of the zebrafish genome. GO term analysis was performed with the TopGO (v2.38.1) R tool (Alexa *et al*, 2006), with the default “weight01” algorithm, the Fisher-test and the org.Hs.eg.db (v3.10.0) R database of the genome wide annotation for humans. Genes without any GO annotation were discarded. Two analyses were run: i- U12 (618) vs U2 (19,939) genes; ii- U2 (19,939) vs U12 (618) genes.

To constitute a list of cilium-related genes, we opted for the combination of three existing databases, and thus defined a gene as cilium-related if 1) it was part of the Gene_Ontology (GO=Cilium), of the Gold_Standard (SysCilia, a curated list of known ciliary components) (van Dam *et al*, 2013) or of the Predicted candidates based on a bayesian integration in CiliaCarta (van Dam *et al*, 2017) (last updated version of March, 18th 2018); 2) it was in the CilDB table v3.0 and had the maximum human ciliary evidence stringency level and at least 3 ciliary evidence (Arnaiz *et al*, 2015, 2009) (last update June 2014); 3) it was a ciliopathy-related gene based on three reviews, (Reiter & Leroux, 2017), (Wheway *et al*, 2019) and (Lovera & Lüders, 2021).

### cDNA library preparation and high-throughput sequencing

One to two micrograms of RNA were sent for RNA-sequencing to IntegraGen Genomics (Evry, France), where a DNA library was generated with the “TruSeq Stranded mRNA Sample Prep” kit (Illumina) that comprises a step of mRNA purification using oligo(dT) beads. RNA-seq experiments have been performed on a HiSeq 4000 sequencer (Illumina), yielding approximately 2670 million of stranded two time 75 bp paired-end reads. Raw RNA-seq data are available upon request. IRFinder v1.2.0 was used to compute the PSI values for U12 introns.

### Cell culture and immunofluorescence

Primary human fibroblasts were cultivated in HAM F10 media (Eurobio, CM1H1000-01) supplemented with 12% fetal bovine serum and 1% penicillin-streptomycin. On Day 1, 4x10^4^ cells were plated onto coverslips in 24-wells plates in order to have them at confluence the next day. On Day 2, cells were starved using HAM F10 media supplemented with 0.5% fetal bovine serum for two days to induce ciliogenesis. For Hedgehog pathway activation, Smoothened Agonist (SAG, 566660, Merck Millipore) was added at 800 nM during the last 24h of serum starvation. Then, cells were fixed in 4% PFA or with cold methanol during 20 minutes at room temperature, and saturated with blocking buffer (PBS 1X, goat serum 10%, bovine serum albumin 1%, Triton 0.1%) for an hour. The following primary antibodies were used: Arl13b (1:500, 17711-1 AP, Proteintech), gamma-tubulin (1:250, ab11316, Abcam), acetylated alpha-tubulin (1:1000, T6793, Sigma) and adenylate cyclase III (1:500, PA5-35382, Invitrogen), diluted in blocking buffer and incubated overnight at 4°C. The following day, coverslips were washed with PBS before being stained with anti-rabbit Alexa 488 (1:1000, A32731, Invitrogen) and anti-mouse Alexa 647 (1:1000, A32728, Invitrogen) for an hour and half, and with DAPI for 10 minutes. Coverslips were finally mounted in FluorPreserveTM Reagent (EMD Millipore, 3457870) and z-stack images were taken using a Zeiss LSM880 confocal microscope. Measurement of the cilium length was performed manually on images of maximum intensity projection of z-stacks, using ImageJ software.

### Zebrafish husbandry and microinjection of embryos

Zebrafish (*Danio rerio*) adults and embryos were maintained at 28.5°C under standard protocols. Breeding and staging were performed as per (Kimmel *et al*, 1995). To avoid pigmentation of embryos dedicated to imaging, 0.003% 1-phenyl-2-thiourea (PTU) was added to embryo medium. The following strains were used: the mixed wild-type ABxTU, and the transgenic lines *Tg(wt1b:EGFP)li1* (Perner *et al*, 2007), *Tg(cmlc2:GFP)* and *Tg(bactin:arl13bGFP)hsc5Tg* (Borovina *et al*, 2010) to label proximal pronephros, heart and cilia, respectively. Live embryos were photographed using a Leica M165FC stereoscope or Zeiss Axiozoom V16. Morpholinos, designed to target the 5’ Stem Loop (5’SL) (5’-GATGTTCTCAGTTAACCTTCATTGA-3’) or the Stem II (SII) (5’-AAACCACCCCCAGACAAGGAAGGTT-3’) regions of u4atac_chr11, were provided by GeneTools, LCC and injected into the yolk at the one-cell stage (quantity ranging from 0.016 to 0.066 pmol (i.e. 135 to 556 pg) per embryo for 5’SL MO, and 0.002 to 0.006 pmol (i.e. 17 to 51 pg) per embryo for SII MO). A five-mismatch morpholino (5’- GATcTTgTCAcTTAACCTTgATTgA-3’) was used as a negative control. For rescue experiments, constructions of human U4atac snRNA of WT or mutated (c.16G>LA, c.51G>LA and c.55G>LA) sequences were previously described (Benoit-Pilven *et al*, 2020). snRNA molecules were synthesized using a PCR amplicon as template (forward primer containing the T7 promoter (in italics): 5’- *TAATACGACTCACTATAGGG*AACCATCCTTTTCTT<u>G</u>GGGTTG-3’ (the underlined G being replaced by A for c.16G>A mutation), reverse primer: 5’- ATTTTTCCAAAAATTGCACCAAAATAAAGC-3’) and the mMESSAGE mMACHINE™ T7 Transcription Kit (ThermoFisher Scientific), followed by a purification using RNA Clean and Concentrator kit (Zymo Research). Seventy-five picograms of snRNA were co-injected with u4atac- targeted morpholinos per embryo.

### RNA extraction, RT-PCR and qRT-PCR

Total RNA was extracted from cultured fibroblasts or a pool of 20 zebrafish embryos and treated with DNase I using NucleoSpin RNA plus kit (Macherey-Nagel). 1.5 μg of total RNA was retro- transcribed into cDNA using random hexamers or oligodT and GoScript Reverse Transcriptase (Promega) following the manufacturer’s protocol. RT-PCR were performed using the GoTaq Green Master Mix (Promega) according to the manufacturer’s instructions (primer sequences in Table S8). PCR products were separated by electrophoresis on 1.5% agarose gel using GelRed nucleic acid gel stain (Biotium). Quantitative real time PCR was performed using the Rotor-gene Q (Qiagen). Reactions were performed in a 20 μl volume, with 0.5 μM of each primer, 10 μl of RotorGene SYBR Green Mix (Qiagen) and 25 ng of cDNA. The amplification was carried out using a denaturation program (95°C for 1 min), 45 cycles of two-steps amplification (95°C for 15s and 60°C for 30s) followed by a melting cycle. Reactions were performed in triplicate, and *ACTB* for fibroblasts or *gapdh* for zebrafish were used as a reference gene (see primer sequences in Table S8). Relative levels of expression of *rnu4atac* genes were calculated using the 2^−ΔΔCt^ method. As for the quantification of intron retention by qRT-PCR, two pairs of primers were designed to target specifically the spliced and unspliced transcripts (see sequences in Table S8). After normalisation to the level of expression of ACTB/*gapdh* gene, the percentage of intron retention was calculated as the ratio of unspliced transcript level to that of the total of unspliced and spliced forms.

### Zebrafish embryo O-Dianisidine staining

O-Dianisidine staining is used to detect hemoglobin in embryos. Staining was performed following a previously described method (Detrich *et al*, 1995). Briefly, embryos were dechorionated at 48 hpf and incubated for 15Lminutes in staining solution containing O-Dianisidine 0.6 mg/mL, 0.01 M sodium acetate (pH 4.5), 0.65% H_2_O_2_, and 40% ethanol. Embryos were post-fixed in 4% PFA for 20 minutes, and dehydrated in methanol before being bathed in a clearing solution of benzyl benzoate/ benzyl alcohol (2:1, vol/vol) for imaging.

### Zebrafish whole mount immunostaining

Embryos were dechorionated, washed twice in PBS, and then fixed in 4% PFA overnight. The next day, embryos were washed in PBS with 0.1% Tween 20 (PBST) and yolks were removed with clean forceps. After incubation for 3 hours in blocking solution PBST+3% BSA, embryos were washed thrice in PBST and incubated overnight at 4°C with anti-acetylated alpha-tubulin antibody in PBST (1:500, T6793, Sigma). The next day, following three PBST washes, samples were incubated with Phalloidin-tetramethylrohdamine B isothiocyanate (1:100, P1951, Sigma) for 20 min at room temperature, washed several times in PBST and incubated overnight at 4°C with anti-mouse secondary antibody in PBST (1:2000, A32728, Invitrogen). Finally, following several PBST washes, embryos were mounted on glass slides using FluoPreserveTM Reagent (Merck) prior to confocal imaging.

### Microscopy of zebrafish embryos

Live and fixed embryos were imaged using Zeiss LSM810 confocal microscope. For live imaging, embryos were mounted in 0.4% low melting agarose mixed with Tricain in glass-bottom Petri dishes (Ibidi). Live beating cilia were imaged by optimizing the signal to noise ratio and the scan speed, as previously described in Borovina *et al*, 2010. Z-stacks were performed to acquire the whole organs (nasal pit, central canal lumen, pronephros lumen, otic vesicle), and quantification of cilium number and range of motion was done manually and blinded on images of maximum intensity projection of z-stacks, using ImageJ software.

### Statistics

All the data are reported as the mean of at least three independent experiments with s.e.m. All hypothesis tests were two-sided, and statistically significant differences (*p*<0.05) were calculated by one-way ANOVA or t-tests as indicated in figure legends. When sample size was too small or when normality was not reached (following Shapiro-Wilk test), a non-parametric test was used. Statistical analyses were performed using GraphPad Prism software.

### Study approval

Animal studies (zebrafish) were conducted in accordance with the European regulations on animal use. Written informed consent was obtained from parents for the genetic study and for the publication of clinical information.

## Data Availability

All data produced in the present work are contained in the manuscript

## Acknowledgments

We thank the families who participated in this study for their contribution to this project. We thank the CBC Biotec biobank for biosample management (Emilie Chopin, Isabelle Rouvet), the PRECI (Plateau de Recherche Expérimentale en Criblage In Vivo) aquatic core facility (Laure Bernard, Robert Renard), the TEFOR facility, and the GENDEV team members for stimulating discussions. This work was supported by CNRS, Inserm, Université de Montpellier, Université Paris 7 and Université Lyon 1 through recurrent funding, the Fondation Maladies Rares (“Small Animal Models and Rare Diseases” program, no. 20161207), the Agence Nationale de la Recherche (no. ANR- 18CE12-0007-01) grants. E.B. was supported by an EMBO long-term fellowship (ALTF-284-2019) and the Novartis Foundation for medical-biological Research (18B112).

