## Supplementary Information and Figures for "Deficiency of the minor spliceosome component U4atac snRNA secondarily results in ciliary defects"

**Supplementary Methods**

*Identification of the zebrafish ortholog of human* RNU4ATAC

Zebrafish has two orthologs for *RNU4ATAC*, on chromosomes 9 and 11, annotated RF00618 in GRCz11. The first gene (chr9, ENSDART00000120294) is located in intron 1 of the *clasp1a* gene, conserving the synteny seen in mammals (human and murine genes reside within *CLASP1* intron 2). However, this gene is transcribed into a 122 bp-long snRNA (compared to 130 bp for human U4atac snRNA) and its sequence identity to human snRNA is 57%. The second gene (chr11, ENSDART00000188971) is transcribed into a 132 bp-long snRNA that shares 68% of identity – with high conservation in functional regions, including a perfect consensus Sm site (AATTTTTGG) that is essential for Sm proteins binding and snRNP stability/function (reviewed in ref.1) (Fig. S2A-C), albeit the synteny is not conserved. Using RNAstructure software, we modelled the structure of zebrafish u4atac/u6atac bi-molecule using one or the other sequence of rnu4atac genes. Human *RNU6ATAC* has two orthologs in zebrafish, on chromosomes 13 and 21, referenced in GRCz11 as RF00619 (Fig. S2A). Both genes are highly conserved; however, u6atac snRNA (chr21, ENSDART00000117190) shares the most of similarity (89%) with the human sequence (Fig. S2A) and the two functional regions are entirely conserved (data not shown). The structure of the zebrafish bi-molecule with u4atac_chr11 is highly resembling that observed in humans, whereas the bi-molecule with u4atac_chr9 looks less relevant (Fig. S2D). In addition, by qRT-PCR analyses, we showed that u4atac_chr9 remained poorly detectable throughout development, while u4atac_chr11 started to rise at dome stage and was strongly expressed at 24 hpf and 48 hpf (Fig. S2E). Finally, further evidence came from the CRISPR/Cas9-mediated knock-out models of u4atac (see below, Fig. S3A). U12 intron retention was only observed in embryos with homozygous deletion of u4atac_chr11, while deletion of u4atac_chr9 had no effects (Fig. S3B). Consequently, contrary to u4atac_chr9 mutant animals which survived until adulthood, a developmental arrest was observed in u4atac_chr11 mutants by 22 hpf (Fig. S3C). Altogether, these data indicated that rnu4atac_chr11 is the sole functional ortholog of human *RNU4ATAC*.

*Generation of zebrafish* rnu4atac *knock-out lines*

*rnu4atac* knock-out lines were generated using CRISPR/Cas9 technology by AMAGEN (France). Two guide RNAs targeting sequences that encompass each of the *rnu4atac* genes (u4atac_chr11: gRNA#1 GAGGCTGAGGGCTGAAATAGC(GGG), gRNA#2, GTGTTAACGGAACATCTATT(TGG); u4atac_chr9: gRNA#1, AAGAGCGTCACAAAACAAAC(AGG), gRNA#2, ATTGGTTAGACAAGCTGGTT(TGG)) were co-injected with Cas9 protein (TACGene, France) into one-cell stage embryos of wild-type AB fish. F0 founders were screened by out-crosses with AB fish and selected based on the germline transmission of the full deletion of rnu4atac to the progeny (LabChip, Perkin Elmer). For u4atac_chr11, F1 animals inheriting a complex sequence rearrangement with two inversions (one of 158 bp, and one of 61 bp) encompassing the deletion of u4atac were selected, whereas, for u4atac_chr9, F1 fish with a 212 bp-deletion repaired by a 7nt insertion (TTGGTTA) were chosen (Fig. S3A). F2 animals issued from out-crosses of F1 fish with AB were raised to adulthood and heterozygous fish were genotyped using a PCR-based method (u4atac_chr11_F: 5’-CCGCTTGGTAACAAGGAAGG-3’, u4atac_chr11_R: 5’-TGTAGTCGTGTCCAAGCAGC-3’; u4atac_chr9_F: 5’-TATTTTCCCAGACACGCCCT-3’, u4atac_chr9_R: 5’-CCGCCCAAAACCTCAACTTT-3’). The phenotype of F3 mutant embryos issued from F2 incrosses were analysed in this study.

*Northern blot analysis*

For northern blot analysis, total RNA was isolated by the TRIzol Reagent method. Briefly, 50 frozen zebrafish embryos were resuspended in 250 mL TRIzol in a 1.5 mL microfuge tube. After homogenization and lysis, 750 μL of TRIzol reagent was added and samples were incubated 10 min at room temperature. Following addition of 0.2 mL of chloroform, vortexing and incubation for 5 minutes at room temperature, the samples were centrifuged at 12,000 x g for 15 minutes at 4°C. The aqueous upper phase, containing the RNA, was transferred into a new microfuge tube. After adding 0,5 mL of isopropanol and incubation at room temperature for 10 minutes, the RNA was recovered by centrifugation at 12,000 x g for 10 minutes at 4°C. The pellet was washed with 75% of cold ethanol and after centrifugation, the RNA was dried and resuspended in RNase-free water. Total RNA samples (10 μg) were separated on a 6% polyacrylamide/8M urea denaturing gel and transferred to HybondN membrane in 0.5x TBE at 40 V for 2 hours at 4°C. After UV treatment, the membrane was prehybridized for 3 hours in 6xSSC, 5xDenhart’s solution, 0,2%SDS at 65°C. Hybridization was performed overnight at room temperature in the same solution with 5’-end-labeled 33P-oligonucleotide, using the following probes: Hs-U4atac, 5’-CACCAAAATAAAGCAAAAGCTGTAG-3’, and Dr-u6atac_chr21, 5’-GTAGGTGGCAATGCCTTAACCATATGTG-3’. Filters were washed twice for 5 min at 37°C in 6xSSC and 0,2% SDS.

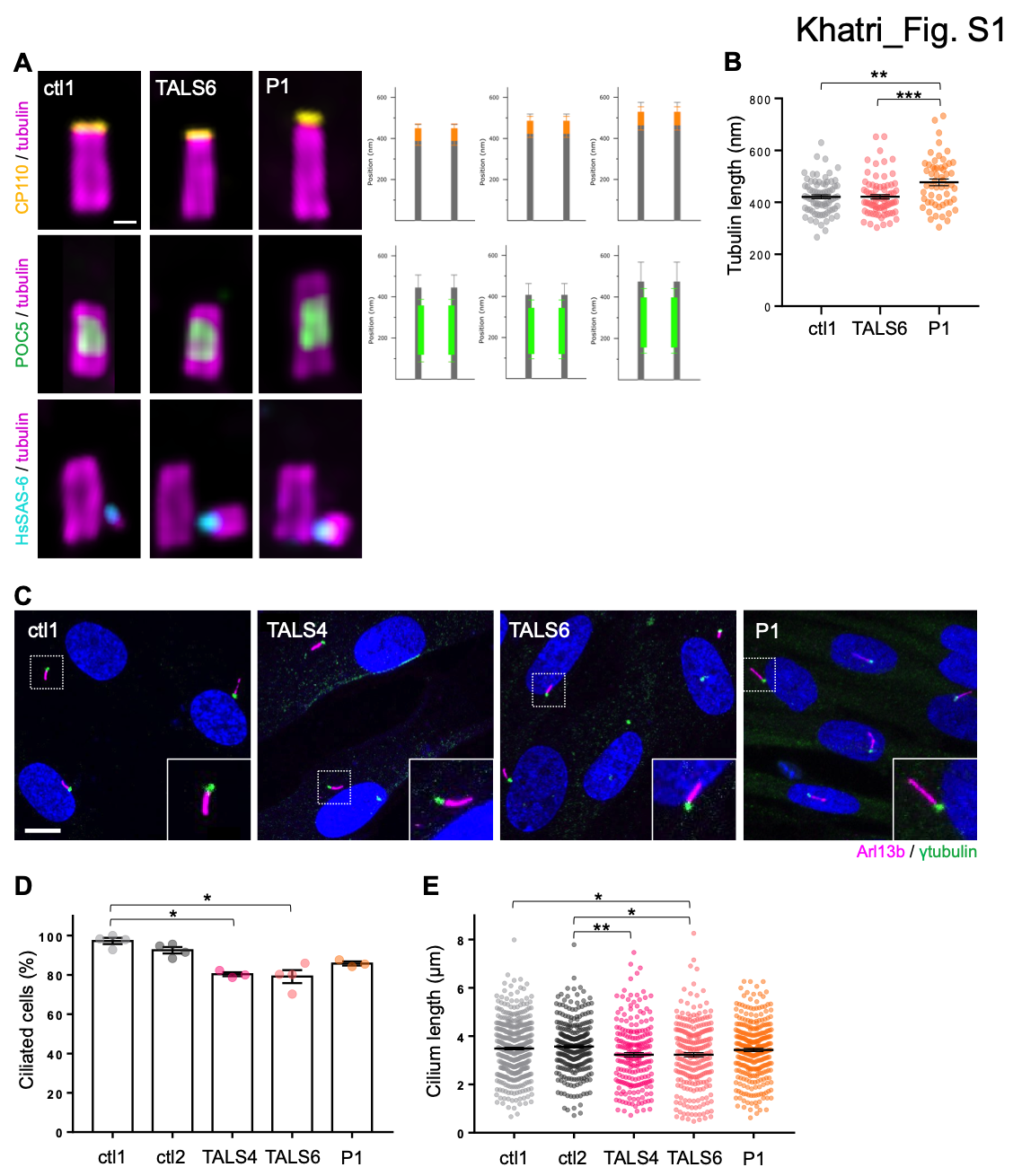

**Figure S1. JBTS/RFMN- and TALS-related *RNU4ATAC* mutations result in moderate ciliary structure alterations**

**(A)** Left panel, representative wild field images of age- and sex-matched control, TALS and JBTS/RFMN patients’ fibroblasts stained for CP110 (yellow), POC5 (green) or HsSAS-6 (cyan) with α/β-tubulin (magenta). Scale bar, 110 nm. Right panel, average position of CP110 and POC5 alongside the centriole of the different cell types. Averages +/- SD from 3 independent experiments (>25 centrioles). Averages +/- SD are as follows, Ctl1 : 391.32 +/- 74.74 to 451.85 +/- 74.74 nm for CP110 and 117.10 +/- 40.06 to 355.28 +/- 62.53 nm for POC5, TALS6 : 421.38 +/- 86.39 to 482.51 +/- 85.69 nm for CP110 and 119.39 +/- 25.73 to 345.94 +/- 64.37 nm for POC5, P1 : 466.90 +/- 100.25 to 531.37 +/- 102.38 nm for CP110 and 154.17 +/- 32.95 to 397.48 +/- 86.99 nm for POC5. **(B)** Length of centrioles quantified using tubulin staining in the three different cell lines. Averages +/- SD from 3 independent experiments (>55 centrioles) are as follows, Ctl : 420.94 +/- 66.90 nm; TALS6 : 421.48 +/- 72.69 nm and P1 : 476.99 +/- 95.75 nm. **p<0.005, ***p<0.001, Kruskal-Wallis test with Dunn’s multiple comparisons test. **(C)** Confocal images of control and patient fibroblasts stained for cilia (Arl13b, magenta) and centrosome (γ-tubulin, green). Scale bar, 10 µm. **(D, E)** Quantification of the percentage of ciliated cells (D) and cilium length (E) seen in (C). Graphs show the mean ± s.e.m. of 3 independent experiments (total number of analysed cells >200). *p<0.05, **p<0.01, Kruskal-Wallis test with Dunn’s multiple comparisons test.

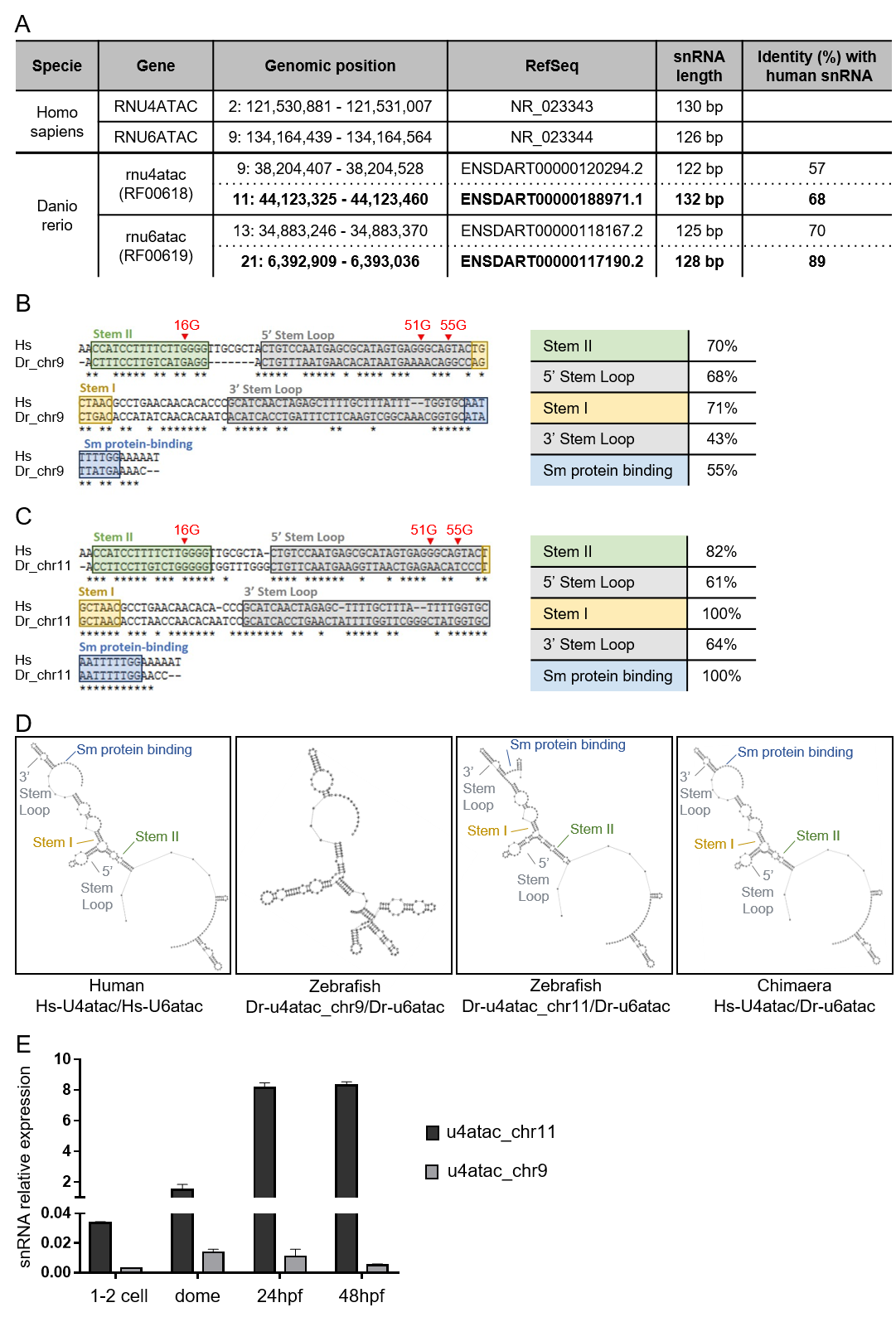

**Figure S2. Characterization of zebrafish u4atac orthologs**

**(A)** Table listing the human and zebrafish orthologs of both *RNU4ATAC* and *RNU6ATAC* genes, with their genomic position, RefSeq ID and transcript snRNA length. For each existing *rnu4atac* and *rnu6atac* zebrafish paralog, the identity to human snRNA sequence is indicated. **(B, C)** Left panels are the sequence alignments of human (Hs) U4atac and zebrafish (Dr) u4atac snRNA (u4atac_chr9 in B and u4atac_chr11 in C), with highlighted in boxes the functional and structural regions of the molecule, by a red arrow the nucleotides of interest (16G, mutated in JBTS patients; 51G and 55G, mutated in TALS patients) and with stars identical nucleotides. Right panels indicate the percentage of identity of the zebrafish sequence to that of the human for each of the functional regions shown in the left panels. **(D)** *In silico* prediction of the U4atac/U6atac bimolecule structure in human and zebrafish, using one or the other of the u4atac paralog (chr9 or chr11) and the bifold function of RNAstructure. The chimeric molecule associating the human U4atac with the zebrafish u6atac, likely to be formed in rescue experiments, is also shown. **(E)** qRT-PCR analysis of relative expression of both u4atac paralogs, normalized to *gapdh*, during four stages of zebrafish development.

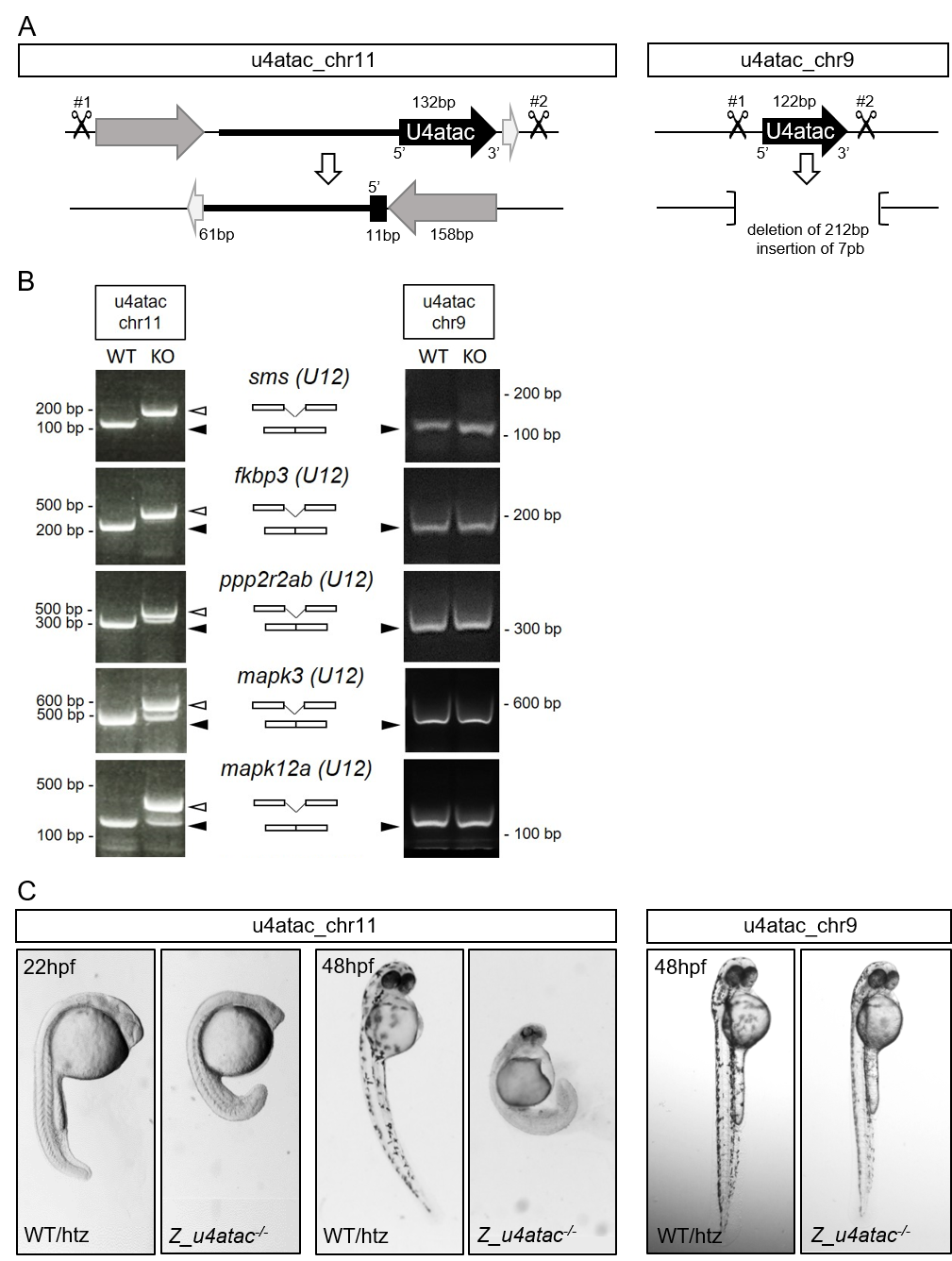

**Figure S3. Generation of CRISPR/Cas9-mediated knock-out zebrafish lines for both u4atac paralogs**

**(A)** Schemas depicting the positions of the two CRISPR guide RNAs used at each locus (chr11 on left panel, chr9 on right panel) to excise the whole u4atac genes. For u4atac_chr11, we obtained an allele with almost a full deletion of u4atac (11bp of 5’ end remained) accompanied by a complex rearrangement with the inversion of two genomic regions, located on either side of u4atac gene. For u4atac_chr9, we selected an allele with a 212 bp-deletion, replaced by a 7 pb insertion (TTGGTTA), that removed the u4atac gene. **(B)** RT-PCR analysis of U12 intron splicing in both u4atac knock-out (KO) lines. Open arrows: transcript with retained intron; filled arrows: spliced transcript. We also tested a U2 intron of *mapk12* gene and we observed its retention in u4atac_chr11 KO line, a finding possibly linked to the fact that 10 splicing factors are encoded by U12 genes. **(C)** Phenotypes of u4atac mutant embryos. For u4atac_chr11 mutant embryos, we observed a delayed development starting at 22 hpf, followed by growth arrest and necrosis. For u4atac_chr9 mutant embryos, no obvious phenotype could be observed at 48 hpf and beyond.

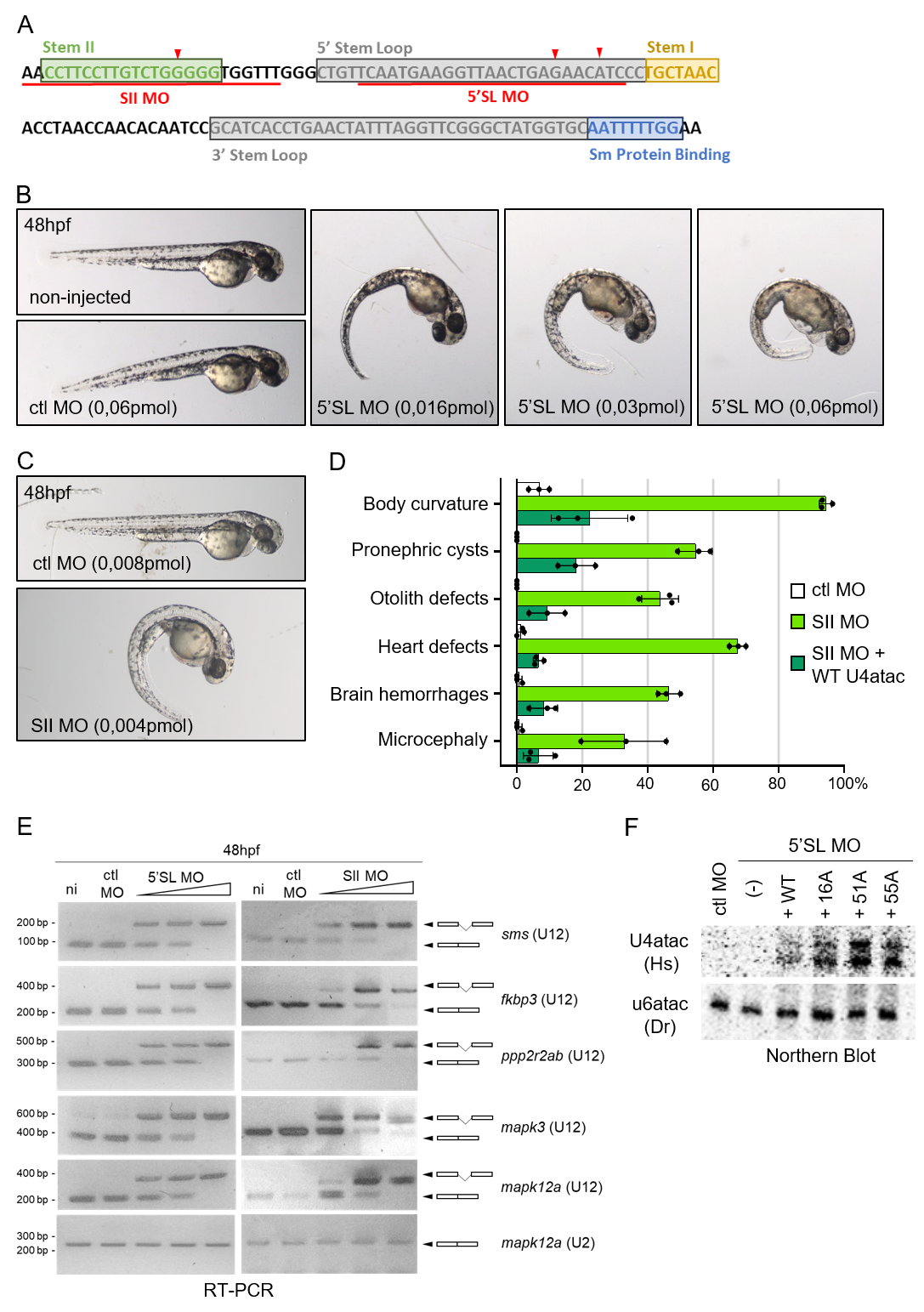

**Figure S4. Characterization of u4atac_chr11 targeting morpholino effects in zebrafish embryos.**

**(A)** Primary sequences of the u4atac_chr11 snRNA with boxes indicating the structural and functional domains. The red underlines show the target sequences of Stem II (SII) and 5’ Stem Loop (5’SL) morpholinos (MO). **(B)** Global morphology of embryos at 48 hpf after injection of control (ctl) or increasing doses of 5’SL MO. **(C, D)** Global morphology of embryos injected with an intermediate dose of SII MO (C), and quantification of embryos (%) displaying one of the various phenotypes observed for SII morphants *versus* control or rescued (SII MO + WT U4atac) animals (D). Graph shows the mean ± s.e.m of three independent experiments (total number of embryos : 150). p<0.05, t-tests comparing SII MO to ctl MO, and SII MO + WT to SII MO. The figures are very close to those obtained with the 5’SL MO (Fig. 4G). **(E)** RT-PCR analysis of U12 intron splicing in 48hpf embryos injected with control or increasing doses of 5’SL or SII MO. The U2 intron of *mapk12* gene is not affected by u4atac_chr11 deficiency. **(F)** Northern blot analysis of human (Hs) U4atac snRNAs injected in zebrafish embryos together with 5’SL MO. Endogenous zebrafish (Dr) u6atac snRNA is used as a loading control.

| **Table S1 : Filtered genes in JBTS/RFMN probands** | | | | | | | | | | | | | |
| --- | --- | --- | --- | --- | --- | --- | --- | --- | --- | --- | --- | --- | --- |
| **Patient** | **Genomic position** | **Gene** | **Gene**  **with** **U12**  **intron** | **Gene associated to ciliopathy** | **Refseq Accession** | **nt change^a^** | **aa change** | **Zygosity** | **PP2** | **SIFT** | **CADD** | **Variation** | **GnomAD (v2.1.1)** |
| P1 F1 : II-1 | **2:122,288,471** | **RNU4ATAC**  **(ncRNA)** |  |  | **NR_023343** | **n.16G>A** | **-** | **hom** | **-** | **-** | **19.3** | **rs750325275** | **23/161196 (0 hom)** |
|  | 2:73,680,967 | ALMS1 |  | ALMS | NM_015120 | c.7310C>A | p.S2467* | het | - | - | 42 | No | No |
|  | 4:15,542,618 | CC2D2A |  | JBTS | NM_001080522 | c.2162C>T | p.P721L | het | 0.997 | - | 35 | rs533830027 | 4/155438 (0 hom) |
|  | 11:117,263,829 | CEP164 | ✓ | NPHP | NM_014956 | c.2603A>G | p.Q868R | het | 0.008 | 0.26 | 12 | No | No |
| P2 F2 : II-1 | **2:122,288,471** | **RNU4ATAC**  **(ncRNA)** |  |  | **NR_023343** | **n.16G>A** | **-** | **hom** | **-** | **-** | **19.3** | **rs750325275** | **23/161196 (0 hom)** |
|  | 12:111,072,571 | TCTN1 | ✓ | JBTS | NM_024549 | c.809C>G | p.P270R | het | 0.992 | 0.09 | 24 | No | No |
|  | 16:56,533,694 | BBS2 |  | BBS | NM_031885 | c.1523A>C | p.Q508P | het | 0.007 | 0.11 | 21.8 | rs115328064 | 72/282890 (0 hom) |
|  | 16:27,720,218 | KIAA0556 | ✓ | JBTS | NM_015202 | c.1582C>T | p.R528C | het | 0.001 | 0.55 | - | rs772590211 | 15/250562 (0 hom) |
|  | 6:108,243,120 | SEC63 |  | PCLD | NM_007214 | c.340-7 delTTTTT | - | het | - | - | - | No | No |
|  | 6:121,560,248 | TBC1D32 | ✓ | OFD | NM_152730 | c.2332C>A | p.P778T | het | 1 | 0.16 | 24 | rs201419425 | 31/280194 (0 hom) |
| Abbreviations are as follows: nt, nucleotide; aa, amino acid; PP2, polyphen2 scores; SIFT, Sorting Intolerant From Tolerant scores; CADD, Combined Annotation Dependent Depletion scores; hom, homozygous; het, heterozygous; ncRNA, non coding RNA. ^a^Variants are numbered according to the indicated RefSeq DNA reference sequence, where +1 corresponds to first nucleotide for *RNU4ATAC* or to the A of ATG start translation codon for coding genes. ALMS, Alström syndrome; JBTS, Joubert syndrome; NPHP, nephronophthisis; BBS, Bardet-Biedl syndrome; PCLD, Polycystic liver disease; OFD, Oro-Facial-Digital syndrome. | | | | | | | | | | | | | |

| **Table S8. Primer sequences.** | | | | |
| --- | --- | --- | --- | --- |
| **RT-PCR primer sequences for splicing efficiency.** | | | | |
| **Gene** | **Species** | **Intron** | **Primer F** | **Primer R** |
| sms | D. rerio | U12 (intron 6-7) | GCAGCGGCAAAGAGCACTATGCTG | AATCCTTACCTCGTAACAATCTCC |
| fkbp3 | D. rerio | U12 (intron 2-3) | AGGGTCTGCTGACGATGAGT | CAGGGTTTGTCTTATTTATTGTCG |
| ppp2r2ab | D. rerio | U12 (intron 7-8) | CCTATGGATCTCATGGTGGAG | TGCTTGTCACAAAGTGCTGA |
| mapk3 | D. rerio | U12 (intron 2-3) | AGACCTACTGCCAGCGCACCCTG | AGCCCTCGCAGGATCTGATACAG |
| mapk12a | D. rerio | U12 (intron 8-9) | GACATCTGGTCAGTCGGGTGCATC | TCTTCAGACTGTAGCTTGGCTGTG |
|  |  | U2 (intron 3-4) | GTTATCGGACTTGTGGATGTGTTC | CATCTGATAGACCAGATACTGCAC |
| **qPCR primer sequences to analyse splicing efficiency or global expression.** | | | | |
| **Gene** | **Species** | **Non-spliced transcript (NSP)** | **Spliced transcript (SP)** | **Total transcripts** |
| RABL2A_F | H. sapiens | GATGGCCAGGACACACTCT | ATTTCTCATGGATGGCTTTCA | **-** |
| RABL2A_R | H. sapiens | AGGATCAGCTTCACATACCACT | CCACAAGGATGGTCTTG | **-** |
| TMEM107_F | H. sapiens | CACCACTGGCCTTTTCTGAC | GCACCCAGAGCCTCATCTC | **-** |
| TMEM107_R | H. sapiens | CACTCCCAACGCTCGAATATG | CACTCCCAACGCTCGAATATG | **-** |
| TMEM231_F | H. sapiens | GCCACAGCCAATTCCATCTT | ACCGTCCTGAATGATCCCAAC | **-** |
| TMEM231_R | H. sapiens | ACCCAGGCGAACTTTACCAT | CCAGAATCCTGGCTGATAAGAAA | **-** |
| TCTN1_F | H. sapiens | TGCTGGTGGATGAACAACAG | CAAAAGTTTGAAATTCATTTTCTTCAGGA | - |
| TCTN1_R | H. sapiens | CCAAAAGGGGCCACGTAAT | GGGAGAGCTCCACCCTT | - |
| IFT80_F | H. sapiens | ATCTTGAAACCATTTGACCCTATTTT | CTGGAAGTGGAGTCATGAGA | - |
| IFT80_R | H. sapiens | GTAGTCCAGCCCACACAG | ACACAGCTTACTAATTCTTGATGC | - |
| ACTB_F | H. sapiens | - | - | ATTGGCAATGAGCGGTTC |
| ACTB_R | H. sapiens | - | - | CGTGGATGCCACAGGACT |
| rabl2_F | D. rerio | CTCAAACACATGCATACACACTC | ATTGATGGTGGTGGTGTATTTGTA | - |
| rabl2_R | D. rerio | ATTGATGGTGGTGGTGTATTTGTA | GATTCCTCATGGATGGATATCG | - |
| tmem107I_F | D. rerio | CAACAACCAAGCTCTTCTGTC | CAGAGGTAGAATGGCAGCAAAA | - |
| tmem107I_R | D. rerio | AAGGAAAAGATCCACCAGTAAATG | CCAATAGCTGAAAAGTGACGGT | - |
| tmem231_F | D. rerio | AAGCCTGACGCATTTAACA | CTCTCTGGTGTCATTCCCGT | - |
| tmem231_R | D. rerio | CACGCAAAATCGGGAATT | CAGAAGCCCGGCTGATAATC | - |
| Ift80_F | D. rerio | GTTGCTGAAGGAGCCCAAAC | GGATGCCTGTGTACTCTGTGG | - |
| Ift80_R | D. rerio | CTTTCCGCTCACAGTCTTGG | ATTTGCAGTCTTCTCCTCCTGA | - |
| tctn1_F | D. rerio | TTTGGGGCACATTCCTTGAC | CGGAACAAGAACGGCAAATGG | - |
| tctn1_R | D. rerio | GCAAGCAATCCTGATCCACA | GCAAGCAATCCTGATCCACA | - |
| pde6d_F | D. rerio | GGATGTTGAAAGTTGAAATCAGAA | GAGATTCTGAAGGGGTTTAAACTAA | - |
| pde6d_R | D. rerio | CTTCGTGTTCTACCCCAGG | CTTCGTGTTCTACCCCAGG | - |
| u4atac_chr9_F | D. rerio | - | - | CTTTCCTTGTCATGAGGACTGTTT |
| u4atac_chr9_R | D. rerio | - | - | GCACCGTTTGCCGACTTG |
| u4atac_chr11_F | D. rerio | - | - | TTCCTTCTCTGGGGGTGGTTT |
| u4atac_chr11_R | D. rerio | - | - | AATTGCACCATAGCCCGAAC |
| gapdh_F | D. rerio | - | - | GTTGGTATTAACGGATTCGGTC |
| gapdh_R | D. rerio | - | - | CACTTAATGTTGGCTGGGTC |

**Supplementary Movie S1.** **Heartbeat of a control morpholino-injected embryo at 48hpf.**

The Tg(cmlc2:GFP) embryo injected with control MO was immobilised and heartbeat recorded using time lapse on Zeiss Axiozoom V16. 500 frames = 25 seconds. Scale bar, 50 µm.

**Supplementary Movie S2. Heartbeat of a 5’SL u4atac morpholino-injected embryo at 48hpf.**

The Tg(cmlc2:GFP) embryo injected with 5’SL u4atac MO was immobilised and heartbeat recorded using time lapse on Zeiss Axiozoom V16. 500 frames = 25 seconds. Scale bar, 50 µm.
